# Inferring viral variants and fitness advantages from longitudinal wastewater sequencing data

**DOI:** 10.64898/2026.11.06.26367358

**Authors:** Auguste Rimaite, Niko Beerenwinkel

**Affiliations:** Department of Biosystems Science and Engineering, ETH Zurich, Basel, Switzerland; SIB Swiss Institute of Bioinformatics, Lausanne, Switzerland

## Abstract

During the SARS-CoV-2 pandemic, wastewater-based epidemiology emerged as an effective tool for monitoring infection dynamics. However, wastewater represents a pooled sample of incomplete viral genomic fragments originating from multiple individuals in the catchment area of a wastewater treatment plant, preventing the assembly of complete viral genomes. Existing computational pipelines are therefore limited in detecting new variants that are absent from clinical sequencing data. We developed FitNMF, an integrated viral dynamics and non-negative matrix factorization model that leverages the temporal structure of wastewater sequencing data and the underlying variant competition to jointly infer mutational signatures, relative abundances, and fitness advantages of circulating variants directly from wastewater next-generation sequencing data. Using simulated data as well as wastewater time-series data collected between 2021 and 2024 across six Swiss wastewater treatment plants, we demonstrate FitNMF’s ability to identify and quantify emerging variants *de novo*, offering a powerful tool for real-time variant surveillance in settings with limited clinical sequencing.

## 1 Introduction

Wastewater-based epidemiology is a cost-effective tool for the early detection and surveillance of pathogens, such as SARS-CoV-2, in the population, and for informing timely public health interventions aimed at mitigating disease outbreaks [1]. Due to extensive clinical sequencing during the COVID-19 pandemic, a substantial amount of clinical data was generated, which served as a ground-truth baseline for effective reproduction numbers or variant relative abundances estimated from wastewater data and for comparisons with estimates from individual samples [2, 3, 4, 5]. However, such comprehensive clinical sequencing is not available for other pathogens, nor is it sustainable for SARS-CoV-2, as COVID-19 has transformed into an endemic infectious disease [6]. Moreover, viral variants from asymptomatic individuals or individuals who refrain from testing cannot be captured through clinical surveillance. Wastewater-based surveillance therefore provides a more representative picture of the variants circulating in the population than clinical surveillance does [7, 8]. More recently, genomic sequencing approaches have also proven effective for monitoring variants of other viruses in wastewater, such as respiratory syncytial virus (RSV) and influenza A virus [9, 10, 11].

Viral genome fragments can be extracted from wastewater and quantified using reverse-transcription quantitative and digital PCR (RT-qPCR and RT-dPCR) methods. It has been shown that estimated concentrations of SARS-CoV-2 in sewage over time correlate with the number of reported clinical cases [12, 13]. Wastewater samples are composite mixtures containing multiple viral genomes circulating in a community. Due to the low concentrations, viral RNA fragmentation, and the presence of PCR inhibitors, analyzing viral genomic composition in wastewater samples is a challenging task. Nevertheless, previous studies have demonstrated that the signal of SARS-CoV-2 variants in wastewater can precede their detection in clinical samples [14], and the relative abundances and growth advantages of known SARS-CoV-2 variants can be inferred using wastewater sequencing data [3, 4, 15].

Due to RNA replication errors and genome editing by host cell defense systems, new mutations constantly appear as the virus replicates, and their accumulation can lead to new lineages and variants of concern [16]. Some of the novel variants may evade immune responses, making antibody-based treatments and vaccines less effective and complicating epidemiological tracking if they are not identified by genomic sequencing from clinical samples. Timely identification and characterization of new variants are crucial for developing vaccines and treatments against SARS-CoV-2 and other viruses as well as for future pandemic preparedness.

Wastewater has demonstrated its utility for the detection of cryptic mutations that represent potential circulating cryptic local haplotypes [17, 18]. Yet, existing computational approaches for the reconstruction of full-length genomes of emerging variants remain limited. Amman et al. [19] employed a two-step hierarchical clustering approach (Ward’s method followed by complete-linkage). Another mutation clustering approach was presented by Lefebvre et al. [20]. Zhuang et al. [21] introduced an independent component analysis-based approach that transforms mutation frequencies into independent and linearly separable sources. However, their assumptions that variants form mutually exclusive clusters of mutations or that they evolve independently rarely hold in practice.

Previous studies [22, 23] demonstrated the potential of non-negative matrix factorization (NMF) to recover mutational patterns of viral variants from wastewater sequencing data. NMF relaxes the independence assumption, jointly infers variant signatures and their relative abundances, and yields interpretable, biologically meaningful representations due to its non-negativity constraints. However, the approach of Ellmen et al. [22] does not use the temporal ordering of the samples, requires imputation of missing values before factorization, and depends on a user-specified number of variants. UnMuted [23] imposes temporal smoothness on the variant abundances using a B-splines approach. However, the number of B-spline basis functions is prespecified rather than inferred from the data, potentially restricting the space of representable temporal trajectories. Moreover, it does not explicitly model the evolutionary mechanisms driving changes in variant relative abundances, which limits mechanistic interpretation and forecasting of the variant trajectories.

Here, we present FitNMF, an NMF-based framework to learn the mutational patterns of viral variants directly from noisy, fragmented wastewater sequencing data, without requiring clinical sequencing data. Our approach leverages the time-series nature of the data to impose smoothness constraints on the reconstructed variant relative abundances. FitNMF integrates a mechanistic variant selection dynamics model with NMF: the dynamics model, fitted jointly with the factorization, constrains the variant relative abundances, enabling quantification of growth advantages and real-time detection of emerging variants. FitNMF also robustly handles missing values and enables data integration across multiple wastewater treatment plants (WWTPs).

## 2 Methods

We first introduce a regularized NMF model without a mechanistic constraint, then present the integrated viral selection dynamics and regularized NMF model, FitNMF, followed by hyperparameter selection. Subsequently, we describe data generation, including sample collection, sequencing, and preprocessing.

### 2.1 Non-negative matrix factorization

NMF factorizes the mutation frequency matrix *A* ∈ [0, 1]^*m×n*^, where *m* denotes the number of mutations, *n* the number of samples, and *A*_*ij*_ the relative frequency of mutation *i* in sample *j*, into two non-negative matrices 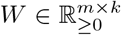 and 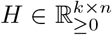, such that *A* ≈ *W H* and *k* represents the number of variants. In this decomposition, each sample (a column of *A*) is approximated as a linear combination of variant signatures (columns of *W*) weighted by their relative abundances (entries of *H*). To quantify the reconstruction error between the observed and approximated frequency matrices, we use the squared Frobenius norm 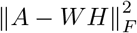.

To allow factorization of matrices with missing values – a common phenomenon due to sequencing dropouts in wastewater data – we imputed missing entries of the matrix *A* with zeros and masked them during loss function evaluation using the binary matrix *M* ∈ {0, 1}^*m×n*^ defined as *M*_*ij*_ = 0 if *A*_*ij*_ is a missing value, and *M*_*ij*_ = 1 otherwise. Then the reconstruction error term in the loss function is 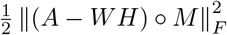, where ° denotes the element-wise (Hadamard) product.

To encourage binary values of the mutational signatures (indicating presence/absence of mutations), we added the regularization term 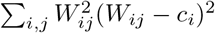 to the loss function, penalizing entries of *W* that deviate from 0 or *c*_*i*_, with the penalty strength controlled by *λ*_*W*_ ≥ 0. When *c*_*i*_ = 1, this term has minima at 0 and 1, irrespective of how much data supports mutation *i*. A mutation *i* whose position is not covered in any sample, or only in a few samples (all or most entries of the row *A*_*i*,:_ are missing), is therefore weakly constrained by the data. Entries of the signature matrix *W*_*i*,:_ for this mutation are driven mainly by the penalty and settle at 0 or 1 depending on whether their random initial value is below or above 0.5. To avoid such unsupported assignments, we scaled the target *c*_*i*_ based on the amount of the data supporting mutation *i* by setting *c*_*i*_ = *n*_*i*_*/*(*n*_*i*_ + *ρ n*_ref_) with constant *ρ* = 0.1, where *n*_*i*_ is the number of sampling days on which the sequence position of mutation *i* was covered (read depth ≥ 30×), and *n*_ref_ is the median of *n*_*i*_ across all observed mutations. A mutation at a never-covered position has *n*_*i*_ = 0 and hence *c*_*i*_ = 0, which forces its entries to zero, whereas a well-covered mutation has *c*_*i*_ ≈ 1. The target *c*_*i*_ reaches the binarization threshold of 0.5 when *n*_*i*_ = *ρ n*_ref_, i.e., when the position is covered in 10% of *n*_ref_. Independent of the *ρ* value, the inferred signatures are further binarized at a fixed binarization threshold of 0.5.

Furthermore, we assume that variant relative abundances change smoothly over time, such that abundances in temporally proximate samples should be similar. We therefore introduce temporal regularization via graph Laplacian smoothing. The node of the graph is a sample, and the edge between samples *i* and *j* is weighted by their temporal proximity, measured by the symmetric Gaussian kernel similarity matrix *K*_*ij*_ = exp{−(*t*_*i*_ − *t*_*j*_)^2^*/β*^2^}, where *t*_*i*_ is the sampling day of sample *i* and *β* is the kernel bandwidth. We normalize *K* by the mean sum across all its rows (or columns),

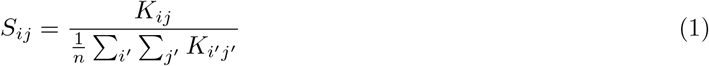

where *n* is the total number of samples, so that the parameter, regularizing the strength of the smoothness constraint, *λ*_*H*_ ≥ 0, has a consistent scale across datasets of different sizes. The graph Laplacian is defined as *L* = *D* − *S*, with the diagonal degree matrix *D, D*_*ii*_ = ∑ _*j*_ *S*_*ij*_. The temporal regularization penalty term is 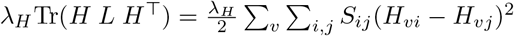, where Tr denotes the trace operator.

The regularized NMF optimization problem is then

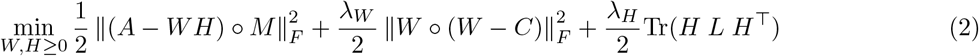

where *C* ∈ [0, 1]^*m×k*^ holds the binarization targets, with *C*_*iv*_ = *c*_*i*_ for all variants *v*.

### 2.2 Integrated viral variant dynamics and NMF model, FitNMF

We assume that the relative abundances of the competing viral variants follow the population dynamics described by the replicator equation 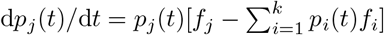 [24]. Here *p*_*j*_(*t*) denotes the relative abundance of variant *j* at time *t*, and *f*_*j*_ its fitness value, assumed to be constant over the observation window.

The solution to the replicator equation is

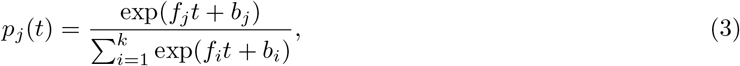

where the offset parameter *b*_*j*_ encodes the log-odds of the abundance of variant *j* relative to the reference variant, at *t* = 0. Since the dynamics depend only on the fitness differences *f*_*j*_ − *f*_*i*_, we fix *f*_1_ = *b*_1_ = 0 for the arbitrary reference variant. For time points *t*_1_, …, *t*_*n*_ and *k* variants, variant relative abundances under the replicator equation model can be described by a matrix *P*(*f, b*; *t*) ∈ [0, 1]^*k×n*^, where *P*(*f, b*; *t*)_*j*:_ = *p*_*j*_(*t*) and *f* ∈ ℝ^*k*^, *b* ∈ ℝ^*k*^, and *t* = *t*_1_, …, *t*_*n*_.

We introduce an additional regularization term to the optimization problem,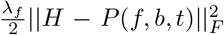, penalizing deviations of the variant relative abundance matrix *H* from the abundances predicted under the replicator model, *P*(*f, b*; *t*), where the strength of the constraint is regulated by the parameter *λ*_*f*_. The variant fitness values *f* and initial condition offsets *b* are inferred jointly with matrices *W* and *H*. Then the optimization problem of the integrated viral dynamics and NMF model, FitNMF, is

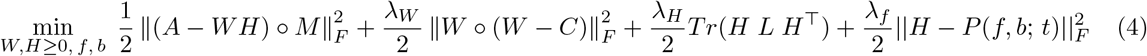

For *λ*_*f*_ = 0, the method reduces to regularized NMF without the mechanistic constraint, and as *λ*_*f*_ increases, *H* is pulled towards the parametric model prediction *P*(*f, b*; *t*). The parameters *W, H*, and (*f, b*) are estimated using the alternating minimization framework (Supplementary Algs. 1, 2). In each iteration, *H* is updated with the model prediction *P*(*f, b*; *t*) held fixed, then *W* is updated, and finally the parameters (*f, b*) of the mechanistic model (Eq. 3) are refitted to the current *H*.

To find the matrices *W* and *H* minimizing Eq. 4 for fixed *f* and *b*, we implemented the multiplicative update algorithm [25]. Each update multiplies the matrix element-wise by a ratio of non-negative terms, which preserves non-negativity. The update rules for *W* and *H* are

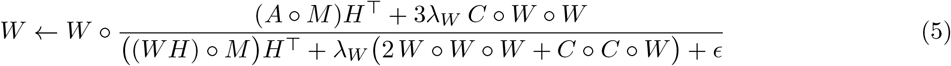

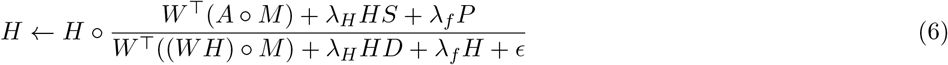

where *ϵ* = 10^−8^ is a small constant for numerical stability. The derivations of Eqs. 5 and 6 are provided in the Supplementary Section S1.3. The fitness coefficients *f* and initial condition offsets *b* are updated by minimizing the objective function of FitNMF with *W* and *H* fixed, using the Broyden–Fletcher–Goldfarb–Shanno (BFGS) algorithm for non-linear gradient-based optimization, with parameters `maxiter:1000` and `gtol:1e-8` in the Python package `scipy.optimize.minimize.` Gradients with respect to *f* and *b* are computed analytically (Supplementary Section S1.2).

These three steps (*W, H*, and (*f, b*) updates) are alternated until convergence, which is evaluated by the relative change in the objective function values between two consecutive iterations, ℒ_*i*_ and ℒ_*i*+1_, normalized by their mean, | ℒ_*i*_ − ℒ_*i*+1_| */*mean(ℒ_*i*_, ℒ_*i*+1_). The process stops when the relative change becomes smaller than the predefined threshold of *τ* = 10^−4^ or when the maximal number of iterations *I*^*max*^ = 5000 is reached. Since the NMF objective is non-convex in (*W, H, f, b*) jointly, convergence to a global optimum is not guaranteed.

Therefore, the algorithm is repeated *n*_run_ = 10 times with random initializations, and the solution with the lowest final objective function value is selected across all runs. During the first warm-up update step, *H* and *W* are each updated once without the mechanistic constraint.

After the final iteration, the entries of the signature matrix *W* are binarized at a threshold of 0.5, to obtain the matrix 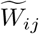 and the relative abundances of the variants are normalized to sum to one for each sample, 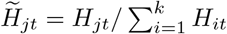. Details of the FitNMF algorithm are provided in Supplementary Section S1.1, Algorithms 1 and 2.

### 2.3 Non-negative matrix factorization with partially known latent factors

Sometimes, several circulating variants are known from clinical data, while others are unknown, for example, because they are circulating only locally or have emerged only recently and therefore are undetected by clinical surveillance. We propose two strategies for incorporating the mutational patterns of known variants into the FitNMF model. First, when the circulating variants are well defined, we include their signatures by initializing selected columns of *W* with their mutation profiles and keep them fixed during the iterations.

While the objective function remains the same as Eq. 4, the update rule for *W* is now

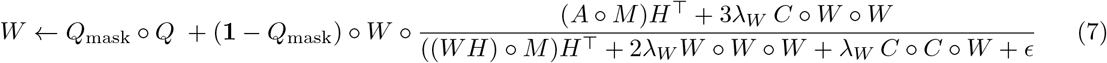

Here, **1** denotes all-ones matrix, and the binary matrix *Q* ∈ {0, 1}^*m×k*^ encodes known variant mutational patterns in its columns (with entries for unknown variants left arbitrary). The logical mask matrix *Q*_mask_ ∈ {0, 1}^*m×k*^ has entries set to one for fixed values and zero for unknown entries. For known variants, values of the signature matrix *W* remain the same as the corresponding masked entries in *Q* during the updates. In contrast, entries corresponding to unknown variants are updated according to the same update rule as in Eq. 5. We note that by setting all entries of matrix *W* to be fixed (i.e., treating every variant’s mutational pattern as known), the method reduces to a regularized deconvolution problem in which only the relative abundances and fitness parameters of the variants are inferred.

Second, when some flexibility is desired, for example, when the signatures of the circulating variants are uncertain due to within-lineage diversity or regional differences, we introduce an additional term weighted by *λ*_*mask*_ ≥ 0, which penalizes deviations of *W* from the reference signatures leading to

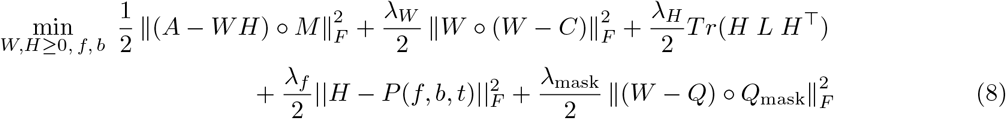

By multiplying by *Q*_mask_, we ensure that deviations from the unknown entries are not penalized in the loss function. The multiplicative update rule for the matrix *W* for masked NMF includes additional terms added to Eq. 5, weighted by the parameter *λ*_*mask*_

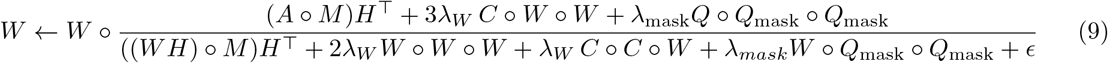

while the update rule for *H* corresponds to Eq. 6.

### 2.4 Integrating data from multiple treatment plants

We extend the FitNMF model to integrate data from multiple locations *z* = 1, …, *Z*, assuming that variant fitness coefficients *f* and signatures *W* are shared between locations, while the relative abundances *H*^*z*^ and offset parameters *b*^*z*^ are allowed to differ. The optimization problem of the multiple-locations integrated viral dynamics and joint NMF approach is

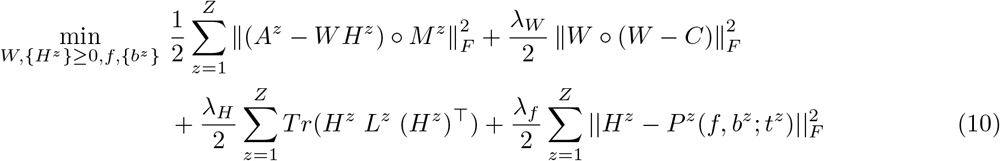

where the entries of *P*^*z*^ are 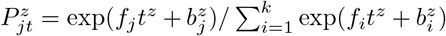 as before. For each location *z*, the time *t*^*z*^ is measured in days since the earliest date across all *Z* locations, and *L*^*z*^ = *D*^*z*^ − *S*^*z*^ is a graph Laplacian built from a Gaussian kernel over that location’s sampling dates. The binarization target is pooled across locations. Its entries *C*_*ij*_ = *c*_*i*_ are determined by the total number of observations of mutation *i* across all *Z* locations. The multiplicative update rules and the gradients for the fitness parameters for Eq. 10 are provided in supplementary section S1.3.1, and the algorithm provided in section S1.1.

### 2.5 Hyperparameter tuning and number of variants selection

To select the hyperparameters *λ*_*W*_, *λ*_*H*_, and *λ*_*f*_, and the number of variants *k*, we employed 10-fold cross-validation (Fig. S6). In each fold *v*, we randomly removed 10% of the observed entries in the data matrix *A*, by treating them as missing to obtain a matrix *A*^*v*^. The binarization target *C* (Section 2.1) was computed once from the complete matrix *A* and kept fixed across folds. We then fitted the FitNMF model using available entries, and computed the reconstruction error of the removed entries (cross-validation error) 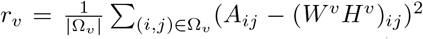, where Ω_*v*_ is the non-overlapping set of removed entries in fold *v*.

We tested multiple combinations (*λ*_*W*_, *λ*_*H*_, *λ*_*f*_) provided in Table S3. For each fold and hyperparameter combination, the model was fitted from multiple random initializations and the solution with the lowest objective value was retained; the settings are listed in Table S4. The average cross-validation error was computed as 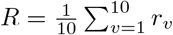, and the parameter combination (*λ*_*W*_, *λ*_*H*_, *λ*_*f*_) with the lowest CV error was selected. This was repeated for *k* ∈ {2, …, 15} (real data) and *k* ∈ {1, …, 6} (simulated data). The number of variants *k* was selected as the last rank before the relative decrease Δ_*i*_ in cross-validation error *R* between consecutive ranks *i* and *i* + 1 first fell below 2%, Δ_*i*_ = (*R*_*i*−1_ − *R*_*i*_)*/R*_*i*−1_, *k* = min{*i*: Δ_*i*+1_ *<* 0.02}.

### 2.6 Wastewater sampling, sequencing, and data preprocessing

The selected time period of SARS-CoV-2 wastewater sequencing data spanned three years, from 1 March 2021 to 1 March 2024. The data were processed using V-pipe [26] with the same configuration as in the Swiss SARS-CoV-2 Sequencing Consortium (S3C) [14] (Section S2.1). We used sequencing data from six WWTPs serving Zurich (996 samples), Altenrhein (990), Laupen (992), Lugano (991), Chur (998), and Geneva (824).

Mutation frequencies were calculated as the ratio of reads carrying the mutated base to the total number of reads covering that position. Coverage and minimal frequency criteria for the filtering are described in detail in Supplementary Section S2.1. After filtering from six location 2,038 mutations were retained. The number of mutations passing the filtering criteria at each sampling location is provided in Section S2.1, Table S2. When factorizing data from a single location, only mutations passing the filter in that location were included, resulting in 1136 mutations for the Zurich WWTP. An overview of the pipeline is shown in Fig. 1.

**Figure 1.**
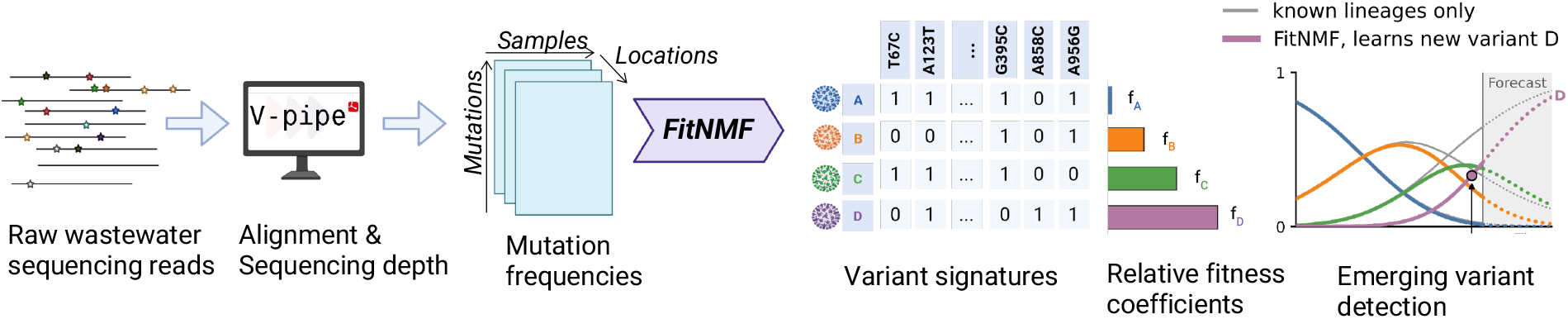
Overview of the pipeline for learning variant signatures *de novo* from wastewater sequencing data using FitNMF. Raw sequencing reads are preprocessed using V-pipe, and the frequency of each mutation is estimated, resulting in a mutation frequency matrix for each location. FitNMF infers variant signatures and relative fitness coefficients. By short-term forecasting of relative abundances, FitNMF can detect the emergence of new unknown variants (variant D). Colors and letters (A–D) denote the inferred variants.

### 2.7 Variant relative abundances and signature mutations

As a ground truth for the set of circulating SARS-CoV-2 variants, we used the variants that were tracked in Swiss wastewater, as reported in https://cov-spectrum.org/stories/wastewater-in-switzerland and https://wise.ethz.ch, and were detected at a relative abundance of ≥ 5% in at least 10 samples in the Zurich WWTP (the largest one in Switzerland). The variants for the deconvolution were selected based on clinical samples, their presence in wastewater tested using COJAC [14] based on unique combinations of the co-occurring mutations. The relative abundances were obtained with the deconvolution tool Lollipop [4] using the following parameters: kernel bandwidth 30, regressor `robust, f scale` 0.01, and `min tol` 10^−3^. SARS-CoV-2 variant signature mutations were obtained by querying Cov-Spectrum [27] (accession date: 2026.08.27) for the mutations present in ≥ 80% of clinical sequences assigned to a particular variant and supported by at least 100 clinical sequences.

## 3 Results

We evaluate the performance of FitNMF, the complete integrated viral dynamics and NMF model that jointly infers mutational signatures, relative abundances, and fitness advantages of circulating variants *de novo*. We applied FitNMF to simulated data and to Swiss wastewater sequencing data collected in 2021–2024.

### 3.1 Application to simulated data

To test the robustness of FitNMF, we simulated datasets that varied in read count overdispersion, fraction of missing values, similarity of ground-truth variants, and temporal dynamics (Supplementary Section S1.6). We first simulated 12 datasets for each of two scenarios with variants evolving under the replicator dynamics model: (*i*) *sequential take-over* of SARS-CoV-2 variants with distinct mutational profiles, and (*ii*) *co-circulation* of four Omicron sublineages with similar mutational profiles (BA.5, BQ.1.1, BA.2.75, and XBB.1.5), with BA.5 not having unique mutations within the comparison set. Each scenario combined three overdispersion levels with four missing-value levels (0, 50, 90, and 95%).

Over both scenarios, the number of variants was correctly inferred for 19/24 datasets (Fig. S7 A, B): for 17/18 datasets with up to 90% missing values, and for 2/6 with 95%. Given the true number of variants, we compared FitNMF with an unregularized baseline NMF (*λ*_*W*_ = *λ*_*H*_ = *λ*_*f*_ = 0), quantifying signature and abundance recovery by the Jaccard similarity *J* and the coefficient of determination *R*^2^ (Section S1.6). The cross-validation-optimal fitness regularization parameter *λ*_*f*_ ranged from 0.4 to 100 (Figs. S8, S10) with a relatively flat optimization surface around the optimal *λ*_*f*_ values (Fig. S9). The smaller *λ*_*f*_ values for the co-circulation scenario and for high fractions of missing data may reflect that the replicator dynamics are harder to recognize when several similar variants co-circulate or the data are extremely sparse.

In the sequential take-over setting (Fig. S11), FitNMF achieved *R*^2^ = 1.00 and *J* ≥ 0.97 at all overdispersion levels, even with 95% missing values, whereas the performance of the baseline model dropped at 95% (*R*^2^ ranged from −0.32 to −0.19, i.e., worse than predicting the mean abundance; *J* from 0.33 to 0.91). In the more difficult co-circulation setting (Fig. S12), the baseline performed poorly even without missing values and little overdispersion (*R*^2^ = 0.39, *J* = 0.68 for BA.2.75), whereas FitNMF reached *R*^2^ ≥ 0.95 and *J* ≥ 0.91 for all datasets with up to 90% missing values. Therefore, missing values degraded the baseline model in both scenarios, whereas FitNMF remained accurate (Figs. S11, S12).

The weekly growth advantage (GA) was estimated relative to a reference variant (B.1.1.7 in the first scenario, BA.5 in the second), 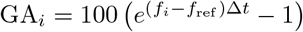 with Δ*t* = 7 days (Supplementary Section S1.4, Fig. S13A,B). This quantity is of epidemiological importance as it allows for short-term forecasting of disease dynamics [28]. The mean absolute error (MAE) between estimated and true GA was 2.9-13.2 percentage points in the first scenario and from 1.8 to 12.7 percentage points in the second, for up to 90% missing values, increasing to up to 38.2 percentage points at 95% missing value level.

We next tested the robustness of FitNMF to rank misspecification in the co-circulation scenario. With a lower rank, FitNMF still recovered the three most abundant variants (signature recovery measured by *J* ranged from 0.9 to 1 in all datasets) but missed the least abundant one, BA.2.75 (Fig. S15). With a higher rank, the most abundant variant was duplicated: its signature was learned twice and its abundance split between the two copies (Fig. S16). All true signatures were still recovered perfectly at 0 and 50% missing values with *J* = 1 for all variants, whereas performance dropped at ≥ 90%. Thus, at low and moderate missing-value levels, a misspecified rank affects only the least or the most abundant variant, and leaves the other variants largely intact. Finally, to test FitNMF when variant dynamics do not follow replicator equation, we simulated two variants evolving by neutral drift (12 datasets; Supplementary Section S1.6). The number of variants was correctly inferred for 9/12 datasets, with errors occurring at high missing-value levels (Fig. S7C). For up to 50% missing values, the cross-validation-optimal *λ*_*f*_ was at most 0.01 (Figs. S8, S10), at least 200-fold below the values selected under replicator dynamics (sequential take-over and co-circulation settings) at the same missing-value levels. Hence, the model found no support for selection-driven dynamics in the data. Under these conditions, FitNMF reconstructed signatures and abundances well (*J* from 0.86 to 1, *R*^2^ from 0.68 to 0.99), whereas the performance of the baseline model degraded (*J* = 0.57, *R*^2^ = −0.45 at 0% missing values) (Fig S17). At 90% missing values, the quality of the reconstructed variants and their abundances by FitNMF dropped (*J* = 0.32, *R*^2^ = 0.25) coinciding with the level at which cross-validation no longer selected *λ*_*f*_ close to zero (optimal *λ*_*f*_ from 0.1 to 80 at 90% and 95%); suggesting that the method could no longer distinguish drift from replicator dynamics at this level of missing values.

### 3.2 Learning *de novo* variant signatures, relative abundances, and growth advantages from SARS-CoV-2 wastewater sequencing data

We applied FitNMF to three years of SARS-CoV-2 wastewater sequencing data from the Zurich WWTP. Ten-fold cross-validation (Section 2.5, Fig. S18) selected *k* = 10 variants and *λ*_*W*_ = 0.01, *λ*_*H*_ = 0.1, *λ*_*f*_ = 5. We matched the inferred signatures to CovSpectrum reference signatures by Jaccard similarity: nine of the ten components corresponded to the dominant circulating variants (Jaccard similarity 0.77–0.94; Fig. 2A,B). The tenth, matched to BA.2.75, had the lowest similarity (*J* = 0.60) and appeared only as sporadic abundance spikes in early 2023.

**Figure 2.**
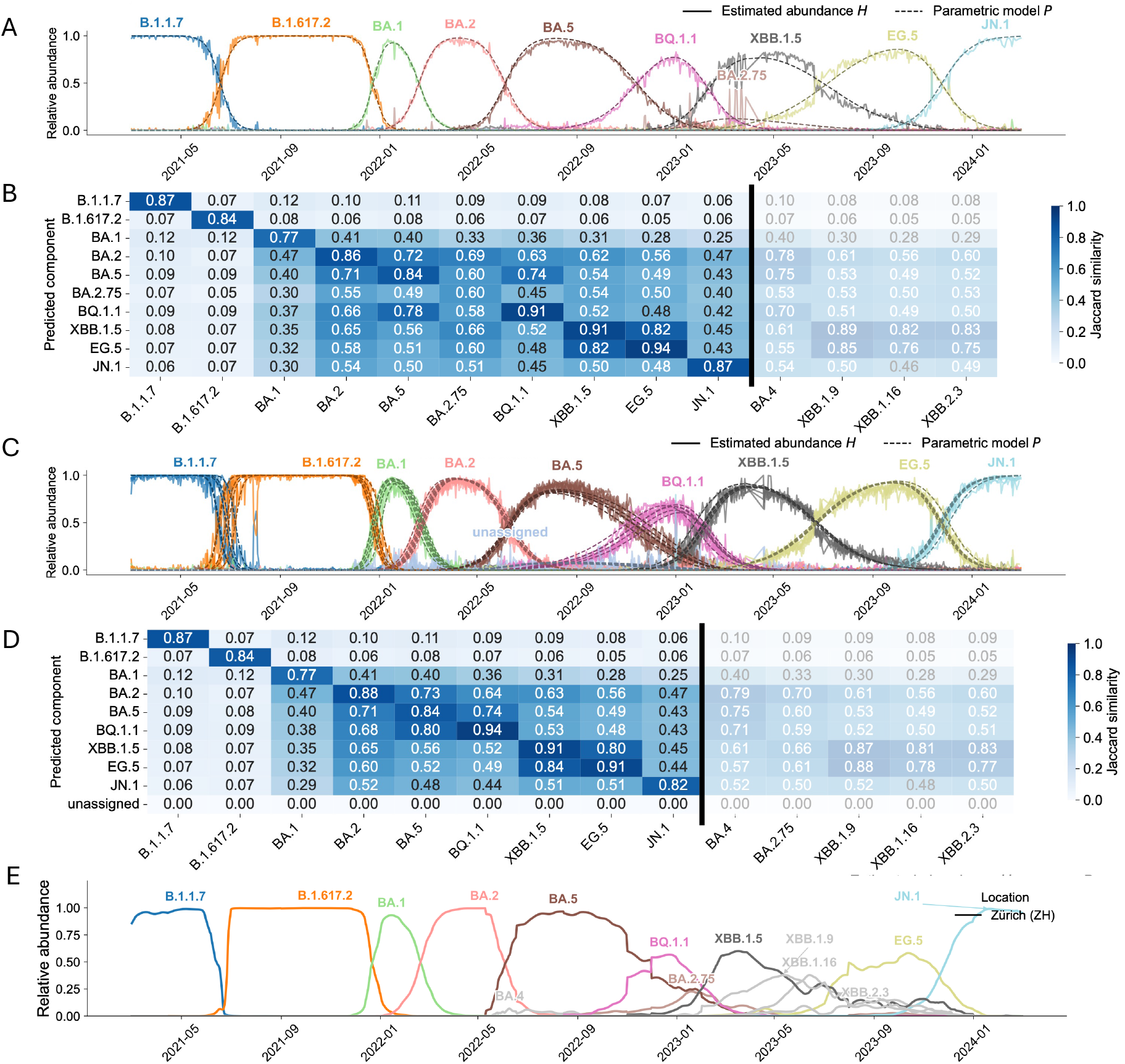
Variant abundances and signatures inferred from SARS-CoV-2 wastewater data, March 2021–March 2024. (A) Estimated relative abundances inferred *de novo* by FitNMF from the Zurich WWTP. Dark dashed lines denote underlying relative abundances predicted under the replicator model. (B) Jaccard similarity between the binarized signatures inferred *de novo* by FitNMF from the Zurich WWTP and the CovSpectrum reference signatures; darker blue indicates higher similarity. Each inferred signature was matched to the CovSpectrum variant signature with the highest Jaccard similarity. CovSpectrum variants not matched by any inferred signature are separated by a black vertical line. (C) As A, from six Swiss WWTPs (Zurich (ZH), Chur (GR), Laupen (BE), Geneva (GE), Altenrhein (SG), Lugano (TI)) jointly. (D) As B, from the joint fit of six WWTPs. (E) Relative abundances from LolliPop deconvolution of the Zurich data with known signatures (variants above 5% for ≥ 10 days; ‘undetermined’ not shown).

The abundances inferred by FitNMF *de novo* agreed with those of the Swiss wastewater surveillance program, obtained with LolliPop where all variants were known from clinical sequencing (Fig. 2A,E): all dominant waves (Alpha, Delta, BA.1, BA.2, BA.5, BQ.1.1, XBB.1.5, EG.5, JN.1) were all identified de novo, and their crossover dates agreed with those of LolliPop. The closely related XBB.1.9, XBB.1.16 and XBB.2.3 that were circulating at lower relative abundances around the same time and whose reference signatures are highly similar to XBB.1.5 (Jaccard similarity 0.82–0.89, Fig. S5) were not resolved into distinct components but captured by the single XBB.1.5 component. BA.4, which peaked below 7%, was not detected, and the component most similar to BA.2.75 appeared only as sporadic spikes in early 2023.

Growth advantages estimated by FitNMF *de novo* aligned well with those estimated by Covvfit using pre-determined variant signatures (Fig. 3). This quantity is of public health interest because it supports short-term forecasting of variant spread [28]. Across 15 co-circulating variant pairs in Zurich, log-scale weekly growth advantages were strongly correlated (Spearman *ρ* = 0.81; variant-label permutation test *p* = 2.8 × 10^−6^; Supplementary Section S1.4, Fig. 3).

**Figure 3.**
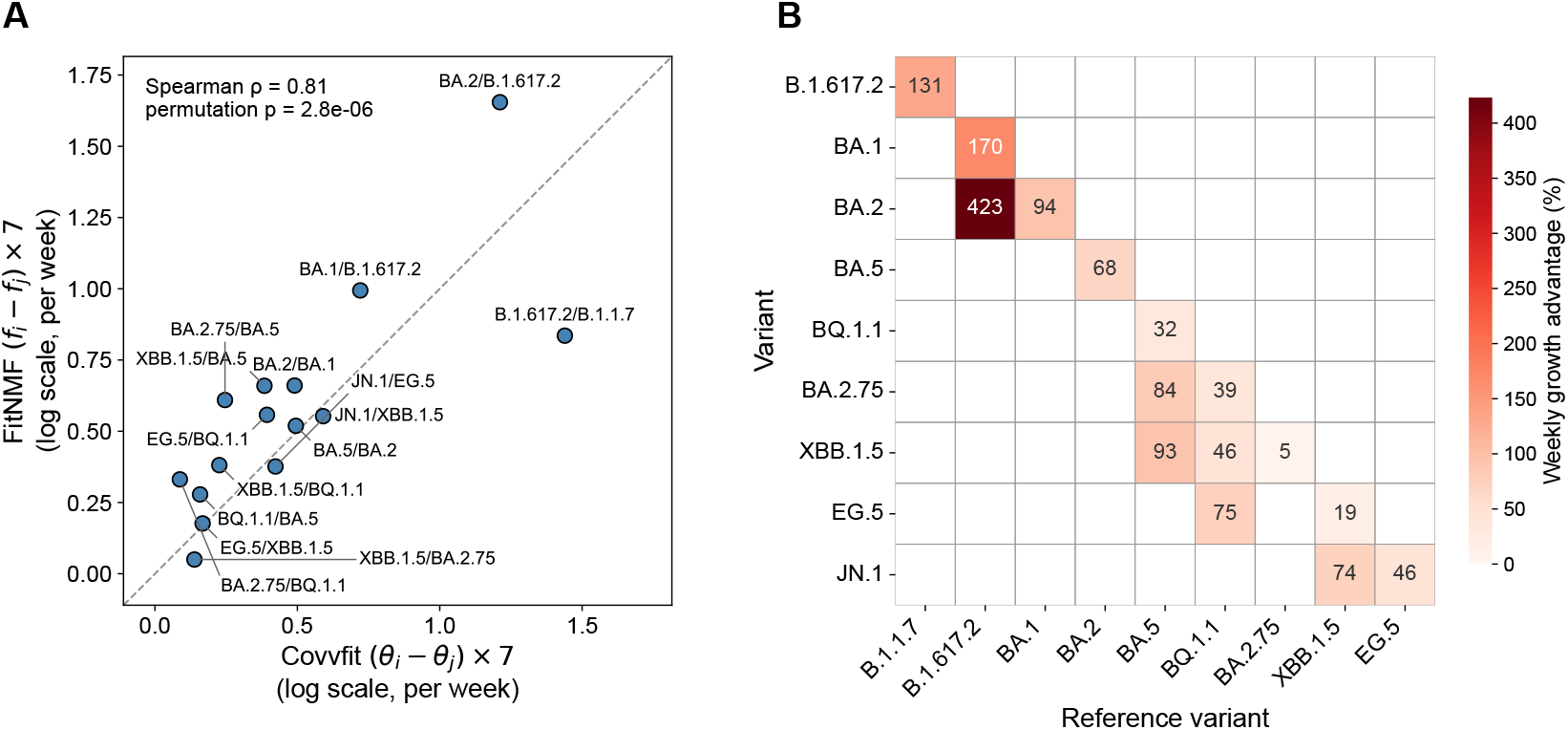
Growth advantages inferred by FitNMF from the Zurich WWTP, 2021–2024. (A) Weekly log growth advantages 7(*f*_*i*_ − *f*_*j*_) of 15 co-circulating variant pairs, estimated by FitNMF versus Covvfit (dashed line: *y* = *x*); Spearman *ρ* = 0.81, permutation *p* = 2.8 × 10^−6^. Only variants matched to a FitNMF component are included. (B) Weekly growth advantage (in %) of the variant in each row relative to the variant in each column, shown for co-circulating pairs only.

Using data from six Swiss WWTPs, FitNMF (Section 2.4), with the same *k* and regularization parameter values as for the Zurich dataset, yielded highly synchronous abundance trajectories across locations, except for the Alpha (B.1.1.7)-to-Delta (B.1.617.2) transition, which differed in timing by about three weeks (Fig. 2C). The tenth component was left unassigned: its binarized signature was empty (Jaccard similarity 0 to all references) and its abundance was low, consistent with fitting noise (Fig. 2D). The predicted signatures were of similar quality to the Zurich fit: across the nine variants assigned in both fits, the mean absolute difference in Jaccard similarity to the CovSpectrum signatures was 0.014 (maximum 0.05).

### 3.3 Detecting an emerging variant using FitNMF

For timely public health interventions, it is important to detect an emerging variant in the population [14]. To test whether FitNMF can detect emerging variants whose signatures are not yet available from clinical sequencing in the real-time settings, we analyzed the interval from March 2021 to March 2023, starting from B.1.1.7 and covering the successive emergence of B.1.617.2, BA.1, BA.2, BA.5, BQ.1.1, and XBB.1.5 (Fig. 4). At each time point, the model was trained on the past observations and used to forecast variant relative abundances 7 days ahead. We compared the regularized deconvolution with fixed signatures of the known variants (Section 2.3) with FitNMF learning the signatures *de novo*, with a rank one higher than the number of known variants. Forecast accuracy was measured by the root mean squared error (RMSE) between predicted and observed mutation frequencies over the forecast window (Supplementary Section S1.5).

**Figure 4.**
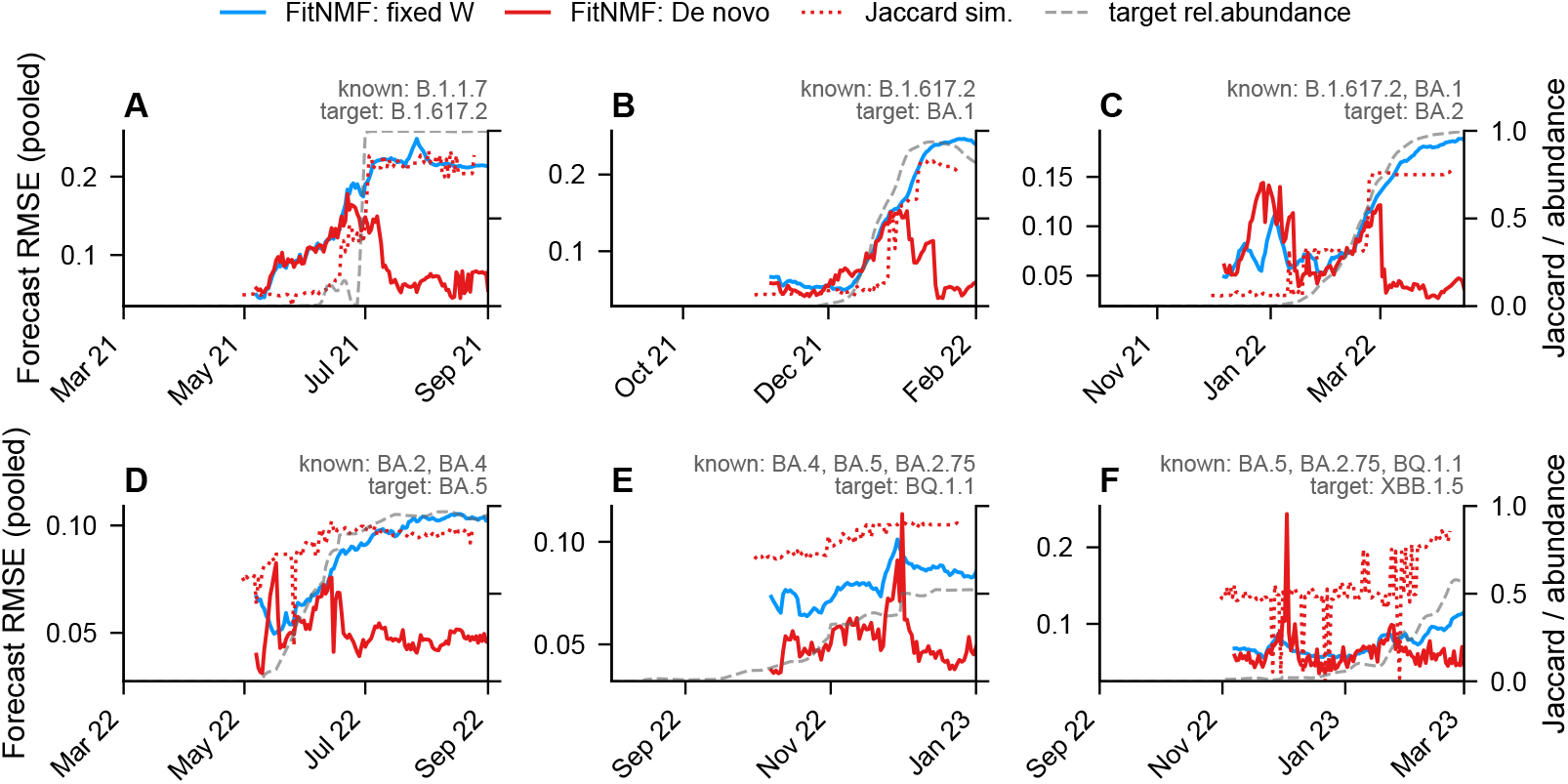
Emerging variant detection using FitNMF. (A–F) The results from SARS-CoV-2 sequencing data from Zurich, March 2021 to March 2023. Forecast RMSE is shown on the left y-axis for FitNMF with fixed, known variant signatures (solid blue lines) and for FitNMF without prior knowledge of the signatures (*de novo*; solid red lines). Each model is trained on all samples up to a given date, and the RMSE measures how well it forecasts the following samples (at least 7 days); it is plotted at the end of this forecast window. Jaccard similarity between the *de novo* component and the CovSpectrum reference signature for the emerging variant is shown on the right y-axis (dotted red lines), plotted at the end of the training window. The grey dashed line denotes the relative abundance of an emerging variant estimated with the Lollipop tool. The known and emerging (target) variants are listed above each panel.

As Delta (B.1.617.2) emerged, the forecast RMSE of the *de novo* model first rose to 0.14 and then declined to 0.048 as the Jaccard similarity between the discovered component and the reference B.1.617.2 signature increased, whereas the RMSE of the fixed model rose to 0.21 and remained high until the end of the period (Fig. 4A). The subsequent intervals (Fig. 4B–F) showed the same pattern: the errors of both models increased as the new variant rose, after which the two models diverged. The divergence was smaller for BQ.1.1 and XBB.1.5, whose signatures overlap more with those of co-circulating variants (Fig. S5). We also evaluated the detection of the emerging XBB.1.5 variant on simulated data in the co-circulation scenario, comparing *de novo*, partially known, and fully known signatures (*λ*_mask_ = 10^3^; Sec. 2.3, Fig. S14). The forecasting errors for *de novo* and partially known signatures diverged from fully known signatures at the same time point, but the model using partially known variants had the lowest forecast error. Overall, by jointly inferring missing variant signatures and fitness parameters and short-term forecasting of relative abundances, FitNMF can indicate the emergence of a new variant.

## 4 Discussion

Wastewater sequencing offers a population-level view of circulating pathogen variants, but most analysis pipelines rely on variant signatures derived from clinical sequencing and therefore cannot characterize variants absent from clinical data. We developed FitNMF, an integrated viral dynamics and non-negative matrix factorization method that jointly infers variant signatures, relative abundances, and relative fitness *de novo* from wastewater time series by constraining abundance dynamics with a replicator model of variant competition.

On real data from the Zurich WWTP, FitNMF recovered the main variant turnovers from Alpha (B.1.1.7) to JN.1, with Jaccard similarity of ≥ 0.6 between inferred and reference signatures for ten variants, and yielded growth advantages that agreed with those of Covvfit [15], which relies on known signatures (Spearman *ρ* = 0.81, permutation *p* = 2.8 × 10^−6^).

The signatures and the fitness parameters are shared across all samples and assumed constant over the observation window. Together they describe the circulating variants, i.e., their mutational signatures and relative fitness, and, through *P*, the abundances expected under the replicator model, and they enable short-term forecasting of relative abundances, which is relevant for public health interventions. In contrast, *H* is allowed to vary across samples, regularized toward *P* and smoothed over time, allowing it to follow deviations from the replicator model and sample-to-sample noise.

Because the strength of the mechanistic constraint is learned from the data, the framework can accommodate variants with different fitness as well as selectively neutral variants whose abundances change by drift. In simulated data without fitness differences between variants and with up to 50% missing values, cross-validation tended to select models with very small values of *λ*_*f*_ giving little to no weight to the parametric constraint. Signatures and abundances were still recovered in this genetic drift case, with the Gaussian kernel smoothing over collection times reducing variance without forcing abundances to follow the replicator model. A weaker constraint may also suit pathogens whose dynamics are not well described by replicator dynamics. With known variants and their mutation signatures fixed, the method reduces to a deconvolution approach that jointly estimates abundances, growth advantages and forecasts future abundances.

The forecasting ability supports detection of emerging variants in the real-time. The forecast errors of a model restricted to known signatures and of a model that learns signatures *de novo* both rise as a new variant emerges, but the error remains elevated only for the known-signature model, whereas it declines for the *de novo* model as the signature of the new variant is recovered.

On simulated data, signatures were recovered with up to 50–95% missing values, with performance depending on variant similarity and abundance dynamics. The fraction of missing values in the real data may vary between datasets; the mutation frequency matrix from the Zurich WWTP contained about 14% missing values over 2021–2024. A joint fit with variant signatures and fitness values shared across the six WWTPs performed comparably to the single location fit in both Jaccard similarity and abundance patterns. Although the timing of the Alpha-to-Delta turnover differed by about three weeks between locations, the variants circulated highly synchronously overall. In total, nine out of ten reconstructed variants corresponded to the ones learned from a single location fit. Since joint-NMF learns factors shared across datasets, a variant that circulates only locally, or location-specific noise, contributes little to the joint loss and may not form a signature of its own.

Our method has several limitations. First, low-abundance variants (BA.4, BA.2.75) were not reliably learned and highly similar sublineages were merged (e.g., several XBB sublineages into XBB.1.5); resolving them may require information beyond mutation-frequency time series, such as mutation co-occurrence within amplicons [14]. Second, fitness is assumed to be constant within the analysis window, which ignores changes in the fitness landscape, for instance due to shifts in population immunity, although this simplification has been shown to yield accurate estimates at lower computational cost [15]. Third, the replicator model is deterministic. In small populations, stochastic fluctuations may make fitness estimates less reliable, although signatures and abundances were still recovered in the neutral-drift simulations. Finally, because the NMF objective is non-convex, multiple random initializations are required. We used 10 initializations, and fewer may reduce the quality of the results. A single fit with fixed hyperparameters is inexpensive, but the cross-validation procedure is computationally more demanding, as its cost grows with the number of hyperparameter combinations, folds, and restarts, and with the size of the dataset. Future work should include uncertainty quantification for the signatures, time-varying fitness and stochastic dynamics models.

In summary, FitNMF identifies and quantifies circulating variants and their fitness advantages *de novo* from wastewater sequencing data and can support viral variant surveillance in settings with limited clinical sequencing.

## Data availability

Raw wastewater sequencing data used in this study is deposited on ENA under project accession number PRJEB44932. The bioinformatics pipeline and all data analysis scripts are available on Github *https://github.com/cbg-ethz/FitNMF*.

## Acknowledgments

We would like to thank all past and present members of the Wastewater-based Infectious Disease Surveillance and Epidemiology (WISE) Consortium. We thank Functional Genomics Center Zurich (FGCZ) for performing the sequencing. We would also like to thank Ivan Topolsky and David Dreifuss for useful discussions at the initial stage of this project.

## Authors contributions

AR (Methodology, Software, Validation, Formal analysis, Investigation, Data Curation, Visualization and Writing – original draft). NB (Conceptualization, Supervision and Funding acquisition). All authors (Writing – review & editing).

## Supplementary Material

## S1 Supplementary

### S1.1 Pseudocode of FitNMF method

Algorithm 2 describes the full FitNMF procedure, which runs *n*_run_ random restarts, selects the one with the lowest final objective, and then binarizes the signatures and normalizes the abundances. Each restart is described in detail in Algorithm 1. Notation follows Section 2. Symbols used in Algorithms 1 and 2 are listed in Table S1.

**Table S1:**
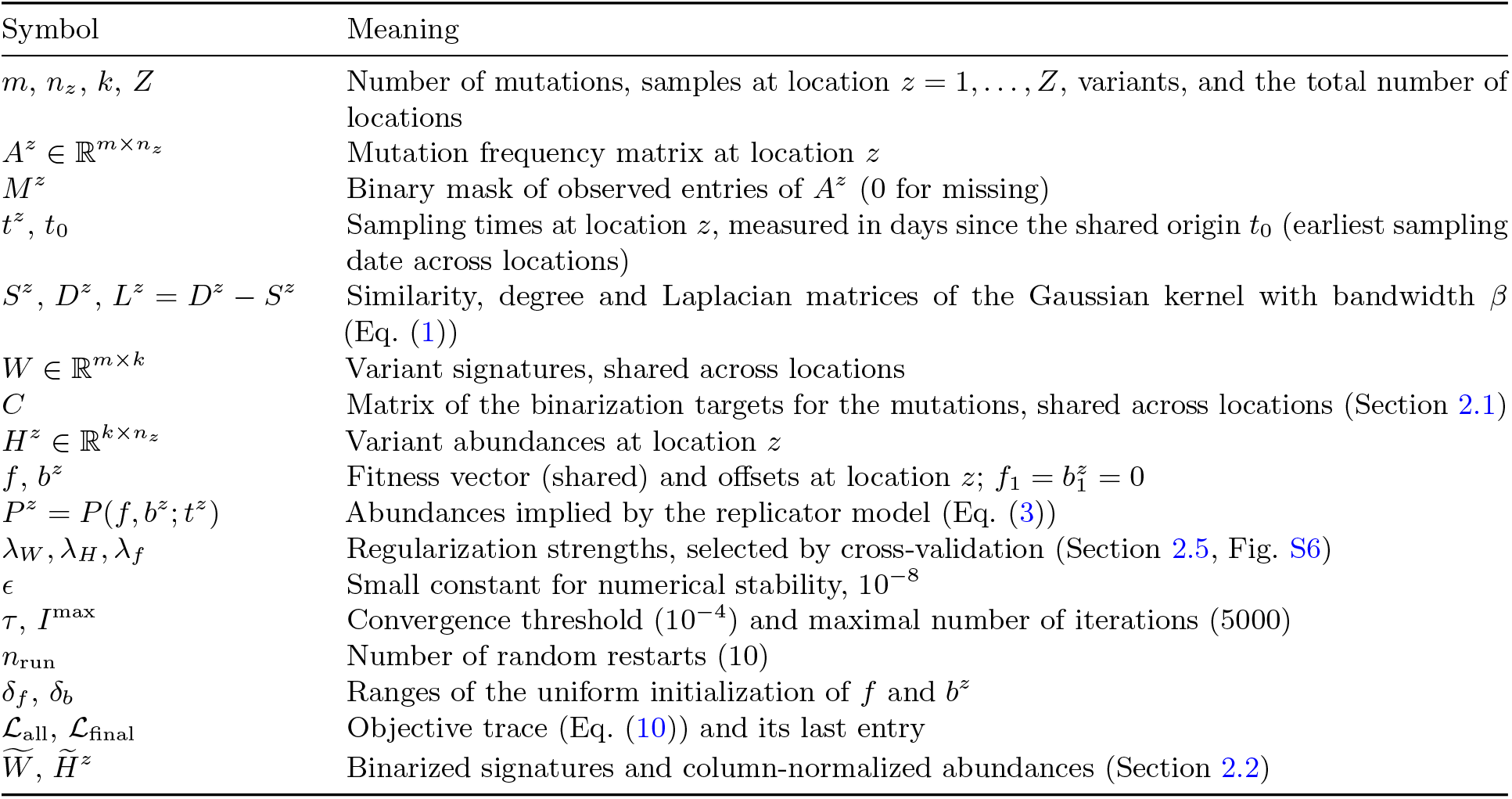
Notation used in the pseudocode.

| Symbol | Meaning |
|---|---|
| m, n<sub>z</sub>, k, Z | Number of mutations, samples at location z = 1, . . . , Z, variants, and the total number of locations |
| A<sup>z</sup> ∈ ℝ<sup>m×n<sub>z</sub></sup> | Mutation frequency matrix at location z |
| M<sup>z</sup> | Binary mask of observed entries of A<sup>z</sup> (0 for missing) |
| t<sup>z</sup>, t<sub>0</sub> | Sampling times at location z, measured in days since the shared origin t<sub>0</sub> (earliest sampling date across locations) |
| S<sup>z</sup>, D<sup>z</sup>, L<sup>z</sup> = D<sup>z</sup> − S<sup>z</sup> | Similarity, degree and Laplacian matrices of the Gaussian kernel with bandwidth β (Eq. (1)) |
| W ∈ ℝ<sup>m×k</sup> | Variant signatures, shared across locations |
| C | Matrix of the binarization targets for the mutations, shared across locations (Section 2.1) |
| H<sup>z</sup> ∈ ℝ<sup>k×n<sub>z</sub></sup> | Variant abundances at location z |
| f, b<sup>z</sup> | Fitness vector (shared) and offsets at location z; f<sub>1</sub> = b<sup>z</sup><sub>1</sub> = 0 |
| P<sup>z</sup> = P(f, b<sup>z</sup>; t<sup>z</sup>) | Abundances implied by the replicator model (Eq. (3)) |
| λ<sub>W</sub>, λ<sub>H</sub>, λ<sub>f</sub> | Regularization strengths, selected by cross-validation (Section 2.5, Fig. S6) |
| ϵ | Small constant for numerical stability, 10<sup>−8</sup> |
| τ, I<sup>max</sup> | Convergence threshold (10<sup>−4</sup>) and maximal number of iterations (5000) |
| n<sub>run</sub> | Number of random restarts (10) |
| δ<sub>f</sub>, δ<sub>b</sub> | Ranges of the uniform initialization of f and b<sup>z</sup> |
| ℒ<sub>all</sub>, ℒ<sub>final</sub> | Objective trace (Eq. (10)) and its last entry |
| W̃, H̃<sup>z</sup> | Binarized signatures and column-normalized abundances (Section 2.2) |

#### Algorithm 1

FitNMF — single restart

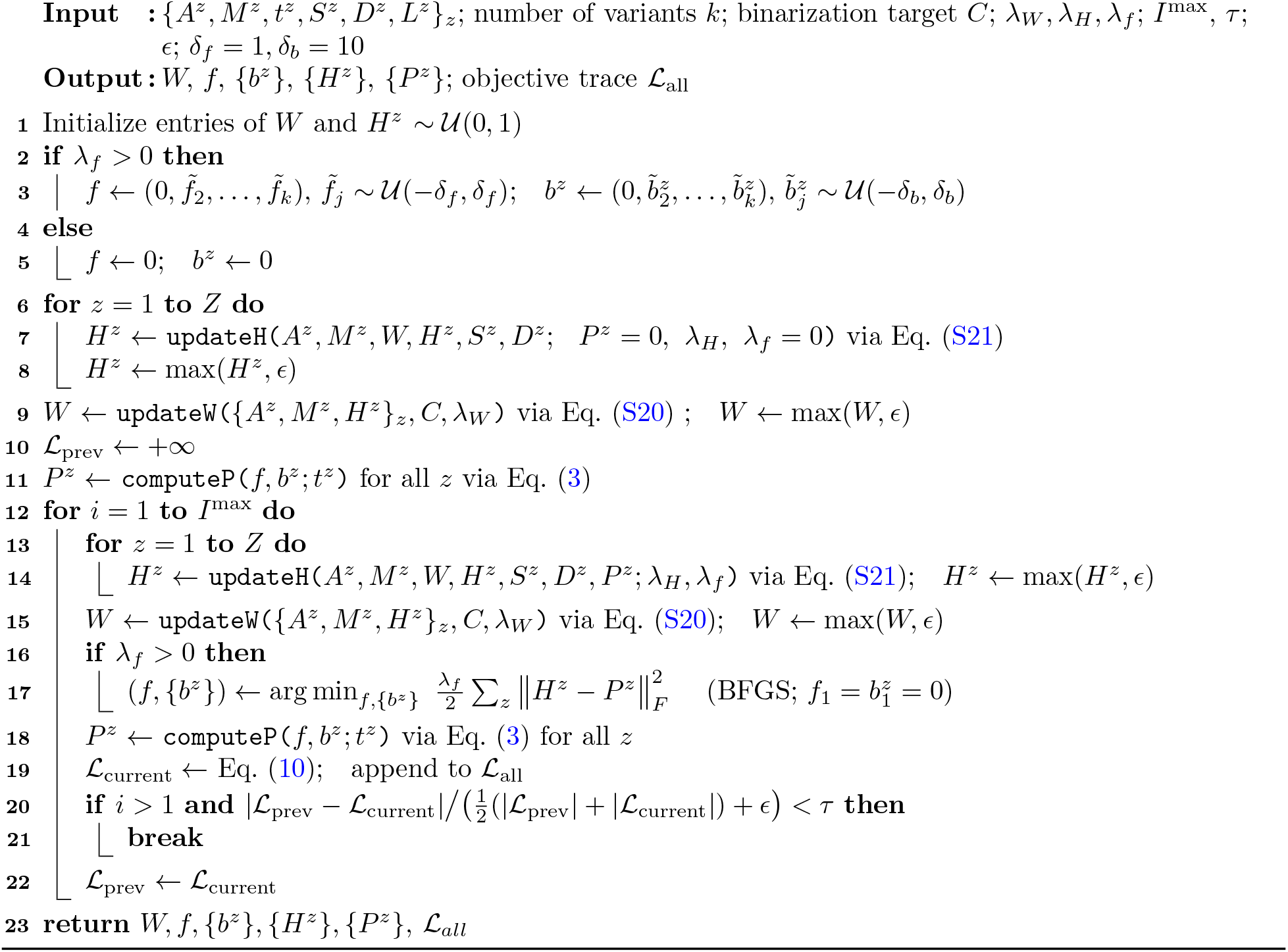

#### Algorithm 2

FitNMF — multi-start

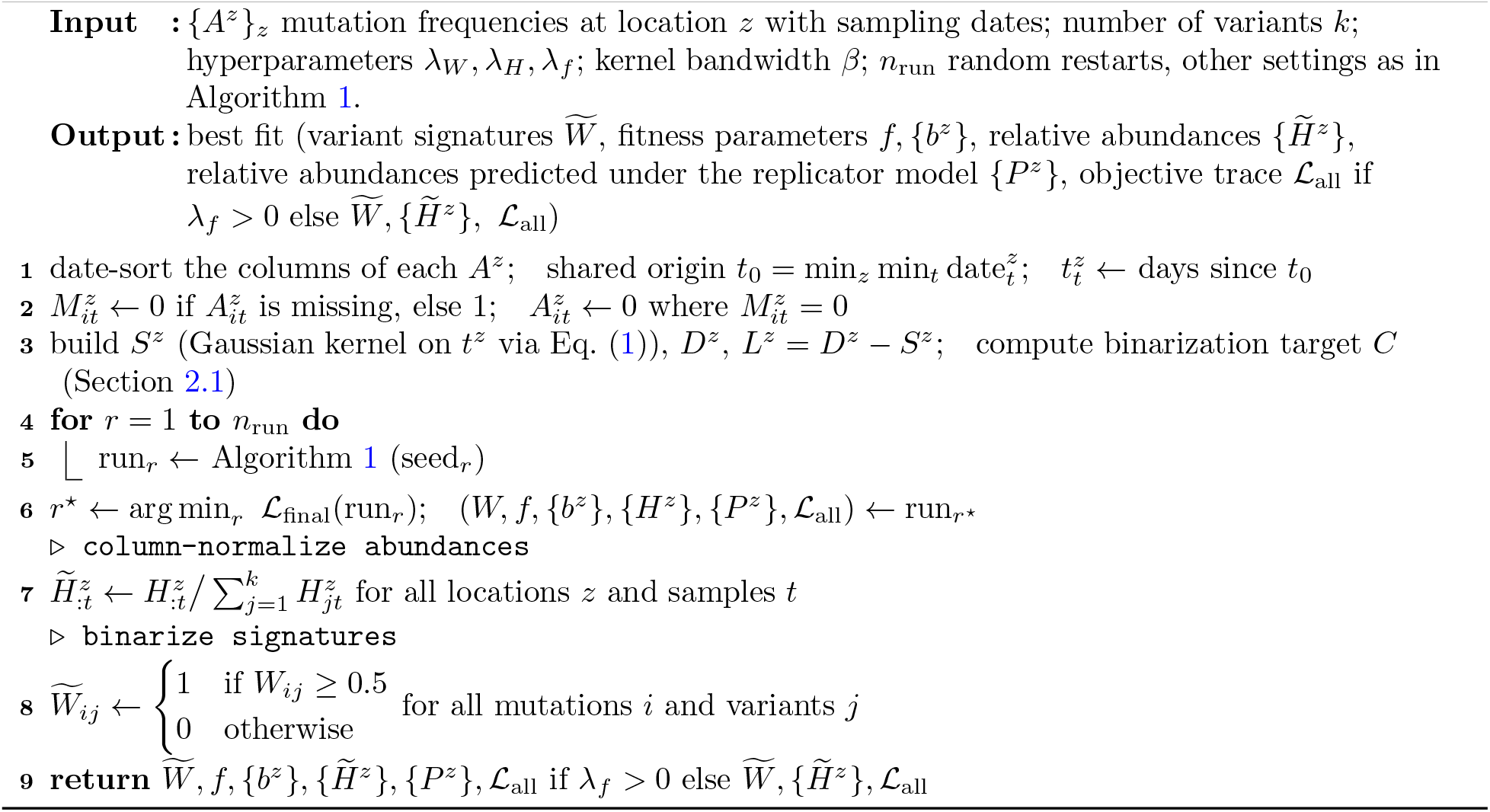

### S1.2 Integrated variant dynamics and NMF model

We modeled variant competition using the replicator equation [24], which describes how the relative abundances of viral variants evolve over time:

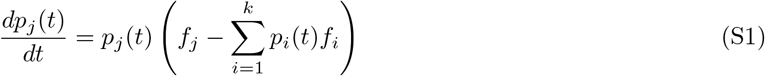

Here *p*_*j*_(*t*) denotes the relative abundance of variant *j* at time *t*, and *f*_*j*_ its fitness value, assumed to be constant over the observation window.

The ODE in Eq. S1 has a closed-form solution, corresponding to a multinomial logistic regression,

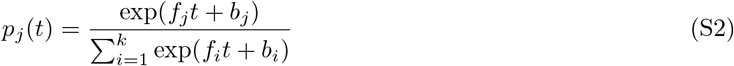

where the offset parameter *b*_*i*_ = log *p*_*i*_(0) − log *p*_1_(0) encodes the log-odds of the abundance of variant *i* relative to the reference variant, at *t* = 0. Since the dynamics depend only on the pairwise fitness differences (*f*_*j*_ − *f*_*i*_), the model is non-identifiable: adding a constant to all fitness values leaves the dynamics unchanged. We resolve this by setting *f*_1_ = 0 and *b*_1_ = 0 for an arbitrary reference variant.

We derive the gradients of the objective function with respect to the fitness parameters *f* and offset parameters *b*, used in the BFGS algorithm (Methods section 2.2). The fitness update step minimizes

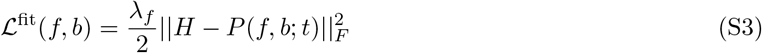

where the entries of the matrix *P* are given by 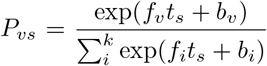 for variant *v* and sample *s*. The derivative of *P*_*vs*_ with respect to the fitness of variant *j, f*_*j*_, is 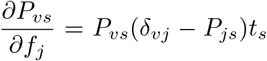, where *δ*_*vj*_ = 1 if *v* = *j*, and *δ*_*vj*_ = 0 otherwise. Similarly, the derivative of *P*_*vs*_ with respect to offset parameter of variant *j, b*_*j*_, is 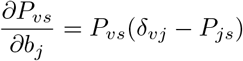.

By the chain rule,

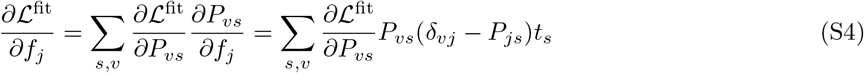

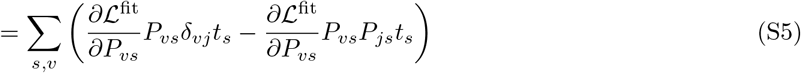

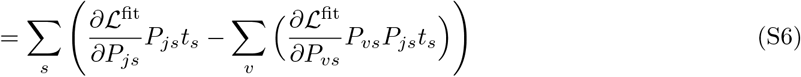

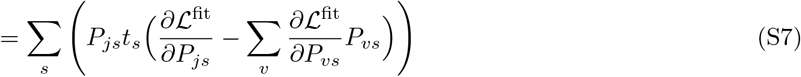

and

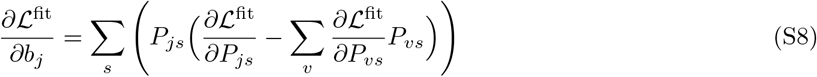

We obtain

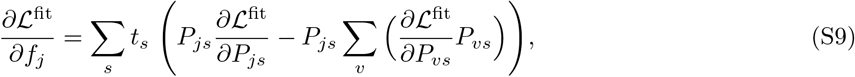

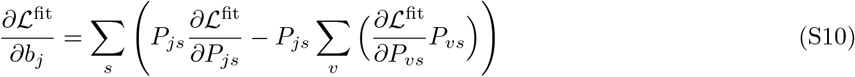

where

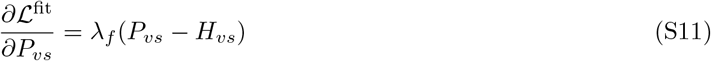

These analytically computed gradients are passed to the BFGS algorithm [29], as implemented in `scipy.optimize.minimize`, jointly updating *f* and *b* while keeping the *W* and *H* matrices fixed during each single step of the alternating optimization algorithm (S1.1).

### S1.3 Derivation of multiplicative update rules

We will first derive the penalty term responsible for the smoothness constraint on variants relative abundances in the loss function in Eq. 2. As defined in Section 2.1, the entries of the symmetric similarity matrix *S* contain normalized Gaussian kernel weights. The diagonal degree matrix *D* of *S* has entries *D*_*ii*_ = Σ_*j*_ *S*_*ij*_. The graph Laplacian is computed as *L* = *D* − *S*.

We penalize weighted squared differences between abundance vectors at nearby time points:

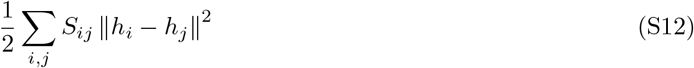

where *h*_*i*_ denotes the *i*-th column of H – the vector of variant abundances at time point *i*. Expanding the square and using the symmetry of *S* gives:

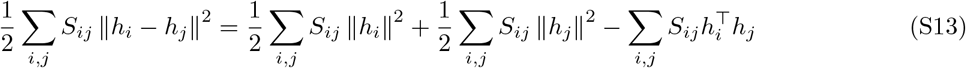

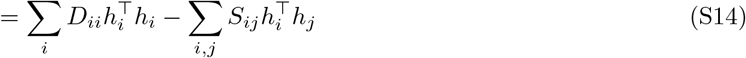

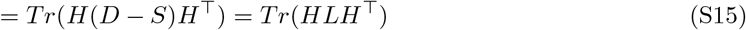

To derive the multiplicative update rules to solve the minimization problem in Eq. 4, we calculate the gradients with respect to *W* and *H*.

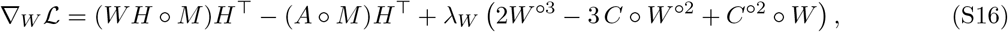

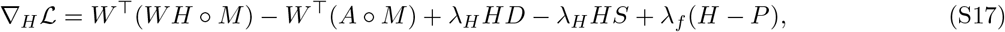

where °*M* denotes element-wise multiplication by matrix *M* and *W* ^°*n*^ denotes element-wise n-th power. Then the update rules for *W* and *H* are

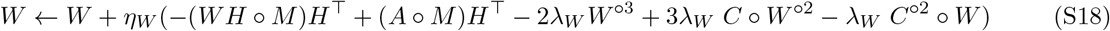

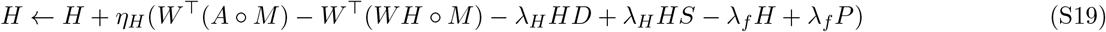

By choosing

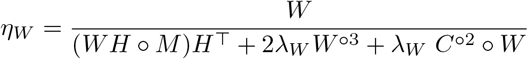

and

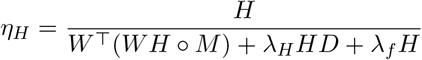

we get multiplicative update rules in 5 and 6.

#### S1.3.1 Integrated viral dynamics and NMF method for multiple locations

The multiplicative update rules for joint-NMF approach (Eq. 10) are provided below.

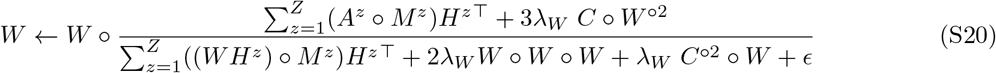

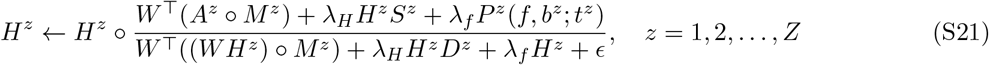

where *ϵ* = 10^−8^ is a small constant added for numerical stability. The fitness update step minimizes the joint loss function across *Z* locations:

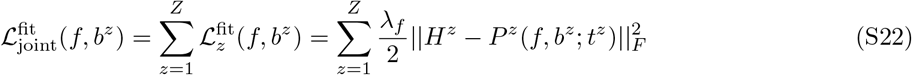

The fitness coefficients *f* are assumed to be shared across locations, while the initial condition offsets *b*^*z*^ are allowed to vary. As in the single-location FitNMF model, the *f* and *b*^*z*^ parameters are updated using the BFGS algorithm. For an arbitrary reference variant we set *f*_1_ = 0 and *b*^*z*^ = 0, where *z* = 1, …, *Z*. Gradients with respect to *f* and *b*^*z*^ were computed analytically (Supplementary Section S1.2). The final expression for gradient with respect to *f* is a sum of the per-location contributions, each evaluated at that location’s own *t*^*z*^. The final expressions of the gradients are:

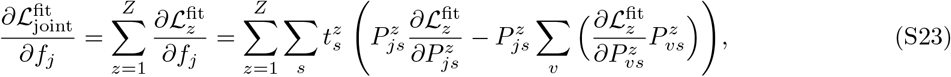

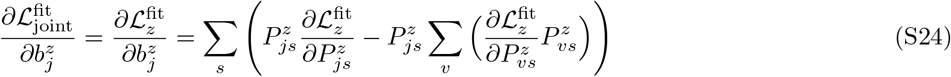

where

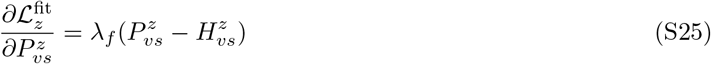

### S1.4 Comparing relative growth advantage estimates against Covvfit

For our model, the relative weekly growth advantages 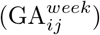 between variants *i* and *j* were calculated as

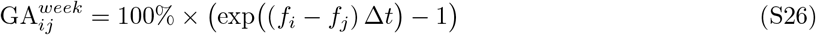

where *f*_*i*_ and *f*_*j*_ are the fitness parameters for variants *i* and *j* obtained from the FitNMF model, and Δ*t* = 7 days converts the per-day fitness difference to a weekly one.

These growth advantages were compared with those obtained from Covvfit [15], a statistical method designed to model the competition of SARS-CoV-2 variants using wastewater-derived sequencing data. Covvfit was run on the relative abundances of SARS-CoV-2 variants obtained with the variant deconvolution tool Lollipop [4], which combines least-squares optimization with kernel-based smoothing for deconvolution. The kernel bandwidth parameter was set to 0.1, regressor was set to `robust, f scale` to 0.01, and `min tol` to 10^−3^, as suggested in [15]. Covvfit was run with 10 random restarts. Samples where “undetermined” fraction of variant relative abundance exceeded 0.01 were excluded from the analysis. Relative weekly fitness differences 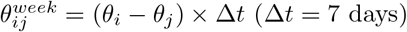, where *θ*_*i*_ and *θ*_*j*_ are the fitness parameters estimated by Covvfit for variants *i* and *j*, were extracted from the Covvfit output (`pairwise fitnesses.csv`) for pairs of variants that co-circulated. Co-circulating pairs were defined from a predefined list of periods in which variants overlapped, and were restricted to variants matched to a component discovered by FitNMF. For each pair we compared 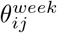 with the FitNMF estimate (*f*_*i*_ − *f*_*j*_) × Δ*t*.

Because pairwise fitness differences are not independent, we assessed their correlation with a permutation test. The fitness coefficients estimated by FitNMF were reshuffled among the *k* variants, the pairwise differences for the same set of pairs were recomputed, and Spearman’s *ρ*_*π*_ with the Covvfit differences was calculated for every permutation. All *k*! permutations were enumerated (*k* = 10, *N* = 10!). The two-sided permutation *p*-value was *p* = *n*_≥_*/N*, where *n*_≥_ is the number of permutations with |*ρ*_*π*_| ≥ |*ρ*_*obs*_|. For both methods, fitness estimates were derived using wastewater data collected in 2021-2024, Zurich WWTP.

### S1.5 Emerging variant detection using FitNMF

We divided the interval into six analysis windows of 5–7 months, partly overlapping, each covering the emergence of a target variant (Fig. 4). Within each analysis window, we used an expanding training window: starting from a minimum length of 60 days, the training period was extended by one day at a time, and at each cutoff the model was re-fitted on all data up to that day. Hyperparameters were selected at each cutoff by cross-validation on the training data only (Supplementary Section S2.3); for simplicity, the same hyperparameters were used for the regularized deconvolution. Variant abundances after the cutoff (*P*_forecast_) were forecast using the parametric replicator equation model (Eq. 3), and the predicted mutation frequencies were calculated as 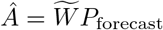, where 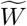 is the binarized mutation signature matrix. For testing, every available sample from the day after the cutoff up to and including the first sample at least 7 days after the cutoff was used. The signature matrix 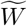 was either (i) fixed to the signatures of the variants known before the emerging target variant, with the rank equal to their number, or (ii) learned *de novo*, without prior knowledge of the signatures, with the rank increased by one. The forecast error was the pooled RMSE over all observed entries of the test samples (Eq. S27):

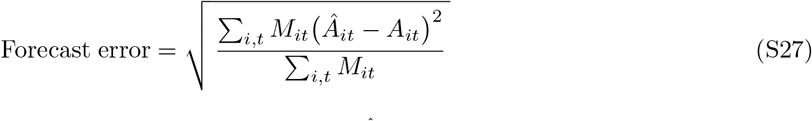

where *A*_*it*_ is the observed frequency of mutation *i* in test sample *t*, Â_*it*_ its forecast, and *M*_*it*_ ∈ {0, 1} indicates whether the entry was observed.

### S1.6 Data simulation

To assess the robustness of the FitNMF method, we simulated data with varying levels of overdispersion, missing values, variant similarity and different temporal dynamics. In the simulated dynamics scenarios (Figs. S1, S3), variant relative abundances followed the multinomial logistic growth model (Eq. S2) or neutral genetic drift. First, we simulated the sequential take-over of genetically different B.1.1.7, B.1.617.2, and BA.1 variants (fitness and offset parameter values: *f*_1_ = 0.0, *f*_2_ = 0.042, *f*_3_ = 0.087 and *b*_1_ = 0.0, *b*_2_ = −5.4, *b*_3_ = −19.0) (Fig. S1A). Second, we simulated a more complex case of multiple Omicron sublineages (BA.5, BA.2.75, BQ.1.1, XBB.1.5) with similar mutational profiles co-circulating (fitness and offset parameter values: *f*_1_ = 0.0, *f*_2_ = 0.052, *f*_3_ = 0.03, *f*_4_ = 0.073 and *b*_1_ = 0.0, *b*_2_ = −8, *b*_3_ = −3.5, *b*_4_ = −12) (Fig. S1B). Variant signature mutations were based on the real data and obtained as described in Section 2.7 (Fig. S2).

To test the model when variant dynamics do not follow replicator equation, we simulated two variants (*A* and *B*) evolving by neutral genetic drift (Wright–Fisher model). Variant *B* started at relative abundance of *p*_0_ = 0.25 in a population of effective size *N*_*e*_ = 500. Every generation (*g* = 3 days), the number of individuals infected with variant *B* was drawn as *X*_*t*+1_ ~ Binomial(*N*_*e*_, *p*_*t*_) and the relative abundance at the next step was estimated as *p*_*t*+1_ = *X*_*t*+1_*/N*_*e*_. Relative abundances were linearly interpolated between generations to obtain daily values. Both variants are defined by 12 unique and 6 shared mutations (Fig. S3).

The sequencing depth *d*_*it*_ of mutation *i* at time point *t* was simulated using Negative Binomial distribution, implemented via the `scipy.stats.nbinom` function in Python:

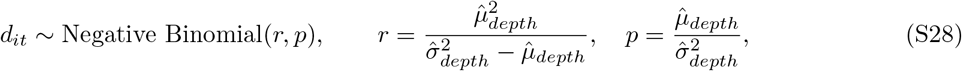

The distribution is defined by two shape parameters *r* and *p*, related to its mean and variance by *µ*_*depth*_ = *r*(1 − *p*)*/p* and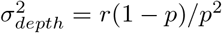. To obtain more realistic depth values, we estimated the mean 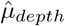 and variance 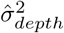 of coverage depth at the variant signature mutation positions with depth *>* 0× across samples collected from March 2021 to March 2024 at Zurich wastewater treatment plant. The estimated values were 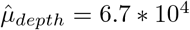,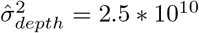.

The true frequency of mutation *i* at time *t, y*_*it*_, is obtained as the product of the mutation signature matrix and the simulated variant relative abundances. Following [4], the sequencing errors were modeled as

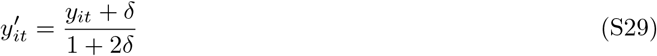

where *δ* parameter (error rate) was fixed to 0.0025. The observed mutation frequencies *m*_*it*_ at mutation *i* and time point *t* were obtained by sampling from a Beta-Binomial distribution, using the `scipy.stats.betabinom` function in Python.

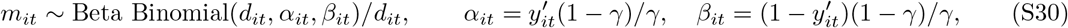

The Beta-Binomial distribution is parameterized by the sequencing depth parameter, *d*_*it*_, and shape parameters 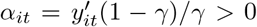, 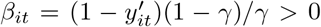, where *γ* is the dispersion parameter (0 *< γ <* 1), controlling the variance around 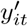(smaller *γ* gives lower variance, approaching a Binomial distribution as *γ* → 0). To test different levels of noise we varied *γ* values (0.001, 0.01, and 0.1). To assess the robustness to coverage dropout, we randomly set 0%, 50%, 90%, 95% of the entries in the simulated datasets to missing values.

The signature recovery was assessed by the Jaccard similarity between the binarized predicted 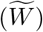 and ground-truth signatures. Since the order of the columns of *W* and the rows of *H* is arbitrary, for the simulated data, we matched the reconstructed and ground-truth variants one-to-one, using the Hungarian algorithm (Python function `scipy.optimize.linear_sum_assignment`), maximizing the Jaccard similarity between the binarized predicted signatures 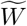 and the ground-truth variant signatures.

The predicted normalized relative abundances 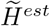 were evaluated by the coefficient of determination:

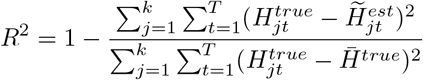

where 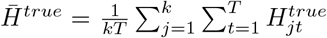 is the mean relative abundance value across all samples (*t* = 1 … *T*) and variants (*j* = 1 … *k*).

**Figure S1:**
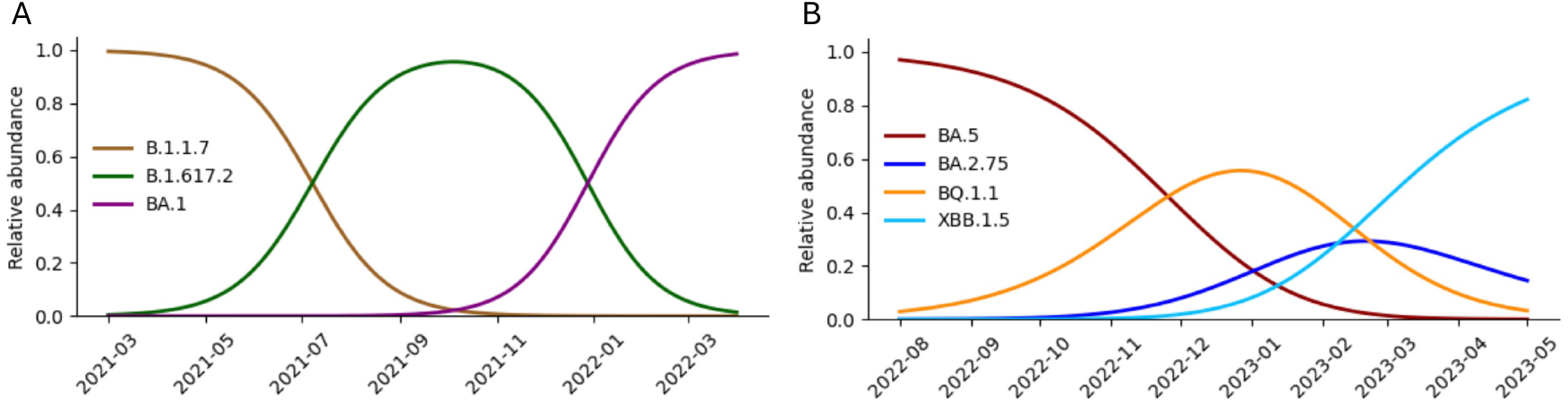
Simulated relative abundances of SARS-CoV-2 variants. (A) Sequential take-over of B.1.1.7, B.1.617.2, and BA.1, variants with distinct mutational profiles. (B) Co-circulation of BA.5, BA.2.75, BQ.1.1, and XBB.1.5 Omicron sublineages with similar mutational profiles.

**Figure S2:**
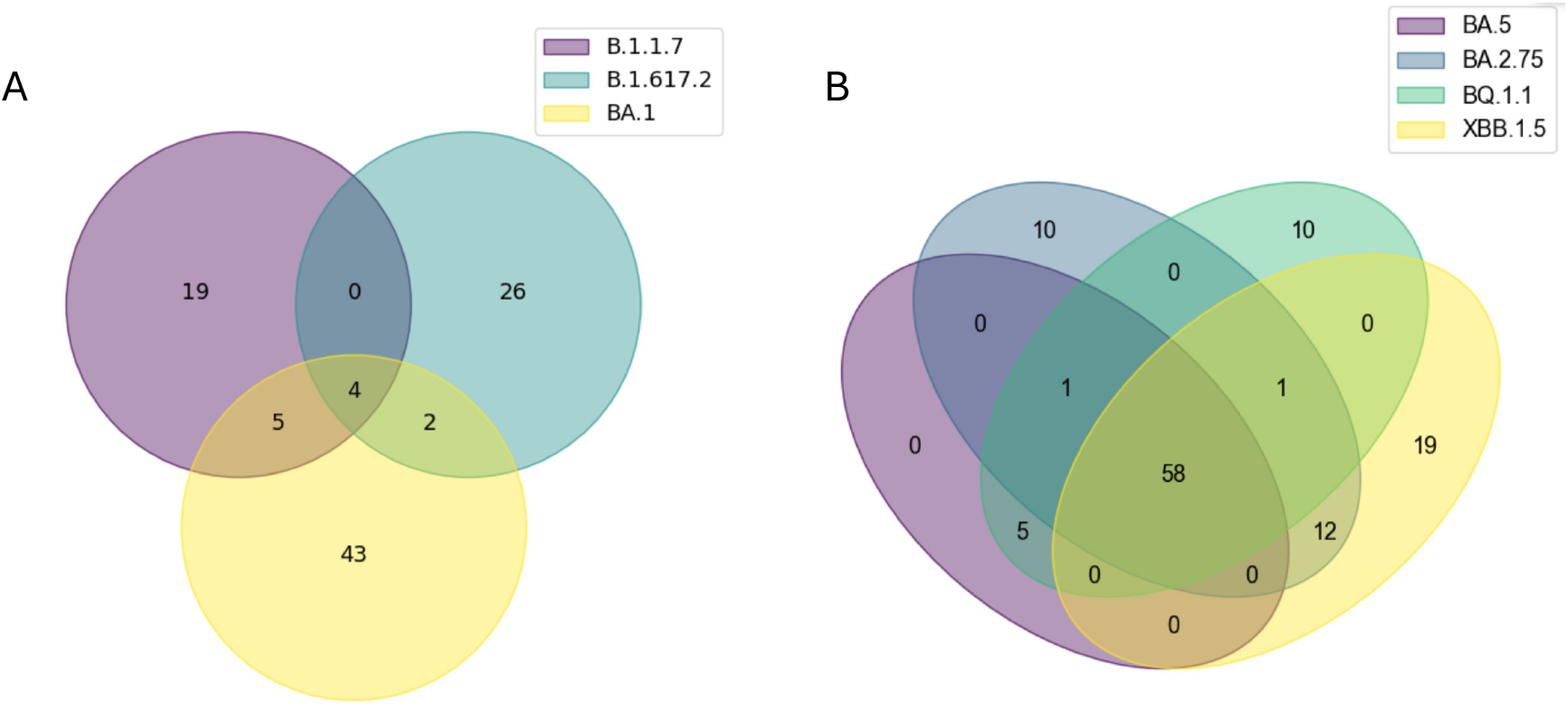
Similarity of mutational profiles between SARS-CoV-2 variants. (A) B.1.1.7, B.1.617.2, and BA.1 variants with distinct mutational profiles. (B) BA.5, BA.2.75, BQ.1.1, and XBB.1.5 Omicron sublineages with similar mutational profiles. Note that BA.5 has no mutations private to it within this comparison set, since all of its signature mutations are also carried by its descendant BQ.1.1.

**Figure S3:**
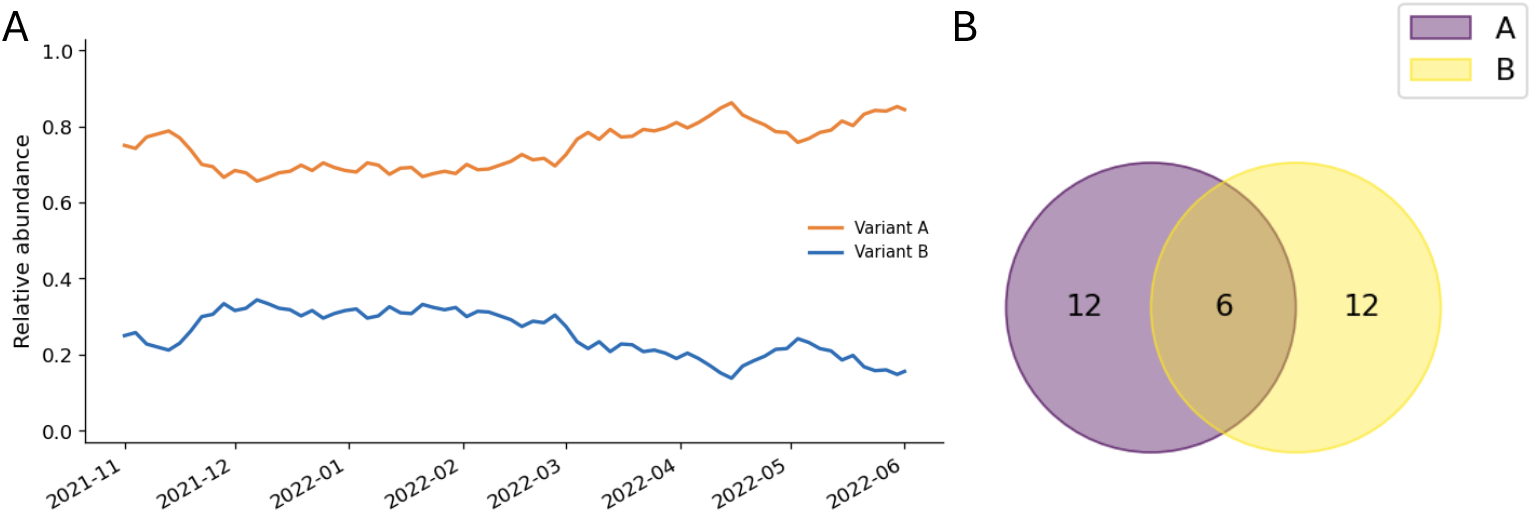
Neutral genetic drift scenario. (A) Simulated relative abundances of two co-circulating variants, A and B. Abundances follow a neutral genetic drift model (*p*_0_ = 0.25, *N*_*e*_ = 500, generation time *g* = 3 days, no fitness difference). (B) Similarity of the mutational patterns of the two variants.

## S2 Supplementary Figures

**Figure S4:**
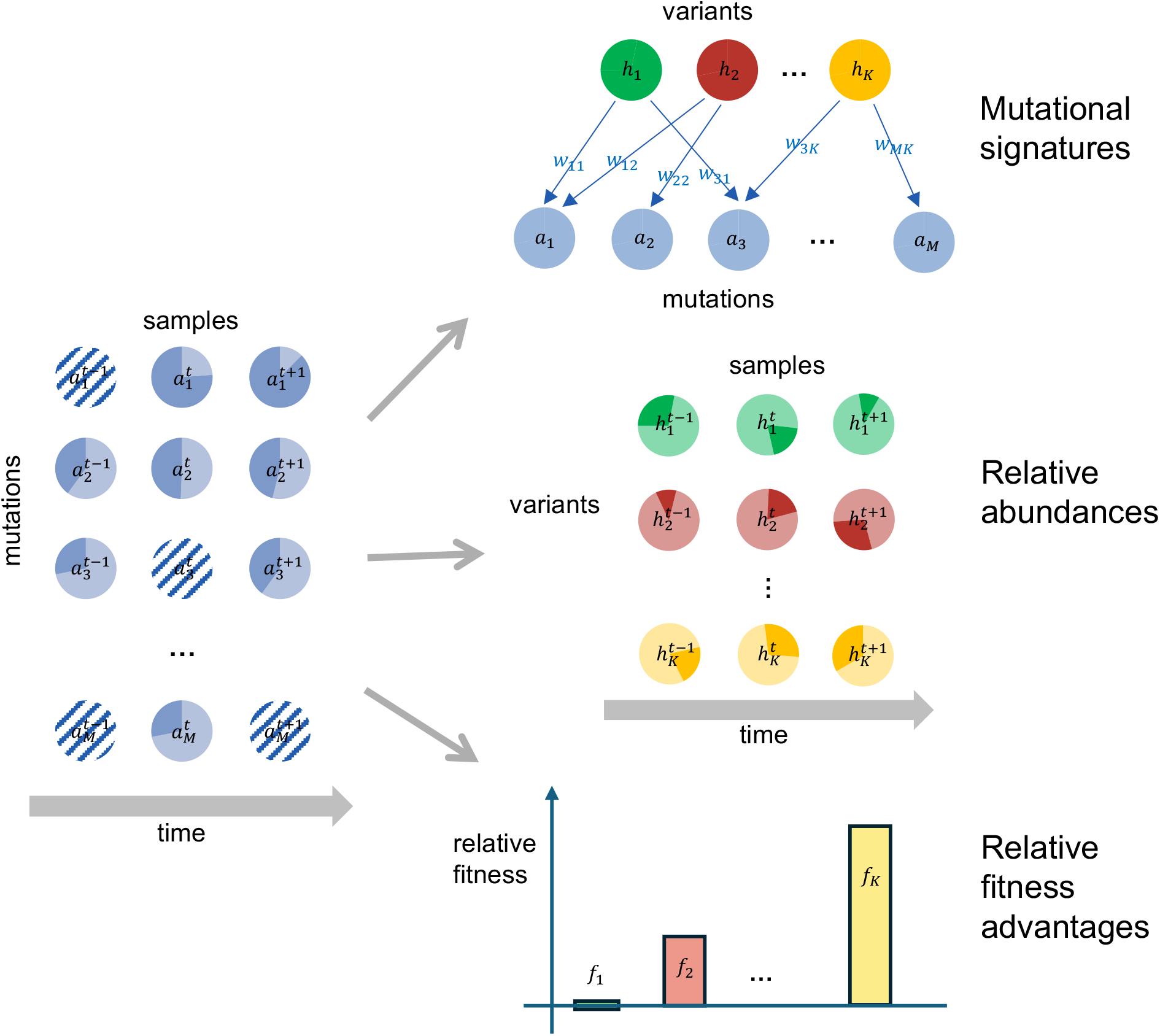
Schematic representation of the FitNMF approach for finding new variants *de novo* from wastewater. Time-series mutation frequency data are factorized into two matrices: variant mutational patterns and variant relative abundances. Different variants are denoted by green, red, and yellow colors. Mutation and variant frequencies are represented as darker shaded portions of the discs. The frequency of mutation *n* at time point *t* is denoted by 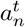, the relative abundance of variant *k* at time point *t* is denoted by 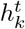, and the assignment of mutation *i* to variant *j* is *w*_*ij*_. The inferred relative fitness coefficients, denoted *f*_*k*_ for variant *k*, are shown as bar plots. Missing values in the mutations frequency data are shown as dashed discs.

### S2.1 Mutation frequency data

The data were processed using V-pipe [26] with the same configuration as in the Swiss SARS-CoV-2 Sequencing Consortium (S3C) [14]. Reads were mapped to the SARS-CoV-2 reference genome NC 045512 (GenBank: MN908947.3) with BWA [30], and per-base read depths were computed from the alignment files using the V-pipe built-in function `aln2basecnt.` Mutation frequencies were calculated as the ratio of reads carrying the mutated base to the total number of reads covering that position. Only single-nucleotide substitutions were used; insertions and deletions were excluded. Frequencies were set to missing where the read depth was below 30x, and to zero where depth was sufficient but no reads supported the mutated base. To reduce the size of the data matrix and to remove likely sequencing errors, mutations were then filtered based on frequency and on the number of sampling days on which they were detected. For joint NMF, a mutation was retained if it reached a frequency of at least 5% on at least 10 sampling days over the three-year period in at least one out of six locations, yielding 2,038 mutations in total. The number of mutations passing these criteria at each sampling location is provided in Table S2. When factorizing data from a single location, only mutations that passed the threshold in that particular location were included.

Reference mutations absent from the data matrix were retained in the evaluation and counted as absent from the inferred signature.

**Table S2:** The number of mutations in real Swiss wastewater sequencing data that passed the filtering criteria at each location and jointly across all locations.

| Location | Mutations that passed the filtering threshold |
|---|---|
| Zurich (ZH) | 1136 |
| Chur (GR) | 1145 |
| Laupen (BE) | 1075 |
| Geneva (GE) | 833 |
| Altenrhein (SG) | 1234 |
| Lugano (TI) | 973 |
| Joint (passed in ≥ 1 location) | 2038 |

### S2.2 Variants similarity

**Figure S5:**
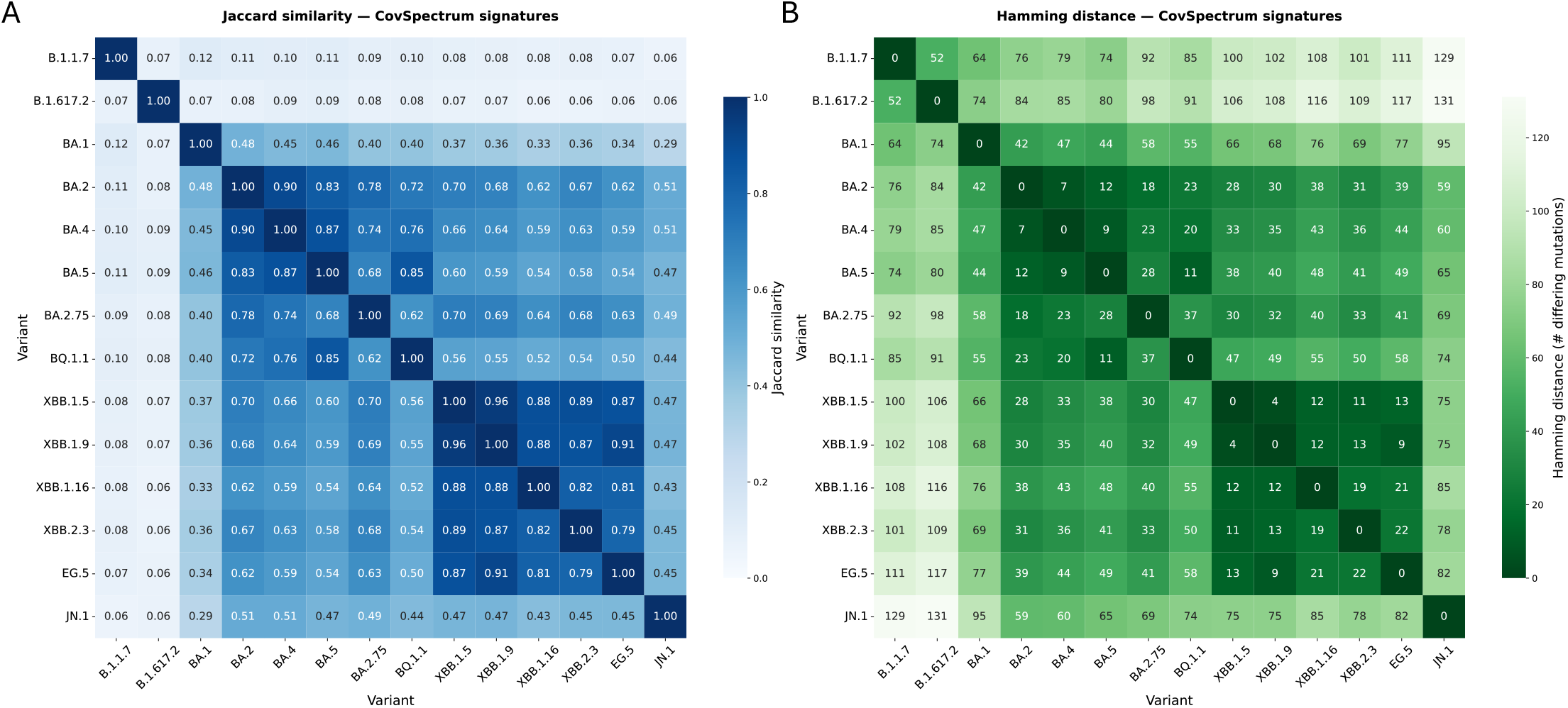
Jaccard similarity (A) and Hamming distance (B) between SARS-CoV-2 variants in real data. Variant signature mutations were obtained by querying Cov-Spectrum [27] (accession date: 2026.08.27) for the mutations present in ≥ 80% of clinical sequences assigned to a particular variant and supported by at least 100 clinical sequences.

### S2.3 Cross-validation

To select the hyperparameters *λ*_*W*_, *λ*_*H*_, and *λ*_*f*_, and the number of variants *k*, we employed 10-fold cross-validation (Fig. S6). We tested multiple combinations (*λ*_*W*_, *λ*_*H*_, *λ*_*f*_) provided in Table S3. For each fold and hyperparameter combination, the model was fitted from multiple random initializations and the solution with the lowest objective value was retained; the settings are listed in Table S4.

**Figure S6:**
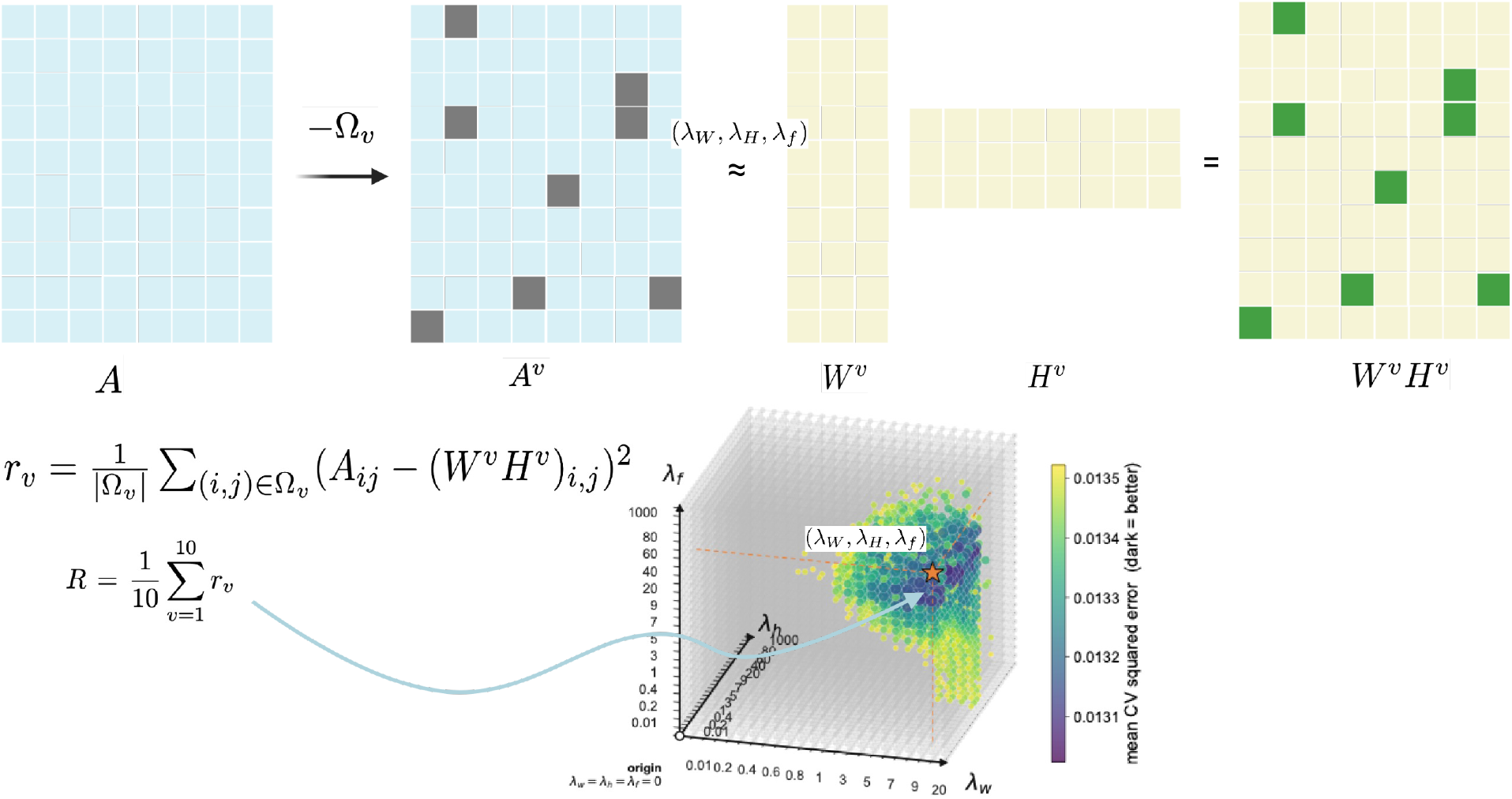
Hyperparameter selection for FitNMF using cross-validation. In each fold, 10% of the observed entries in matrix *A* were held out to obtain matrix *A*^*v*^. We then fitted the NMF model with different values of hyperparameters (*λ*_*W*_, *λ*_*H*_, *λ*_*f*_) and the number of variants *k*. We computed the reconstruction error for the removed entries as 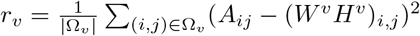, where Ω_*v*_ is the non-overlapping set of masked entries in fold *v*. For each fold and hyperparameter combination, we ran NMF with multiple random initializations, selecting the solution with the lowest objective function value. The average reconstruction error was computed as 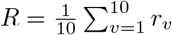, and the combination (*λ*_*W*_, *λ*_*H*_, *λ*_*f*_) minimizing *R* was selected.

**Table S3:**
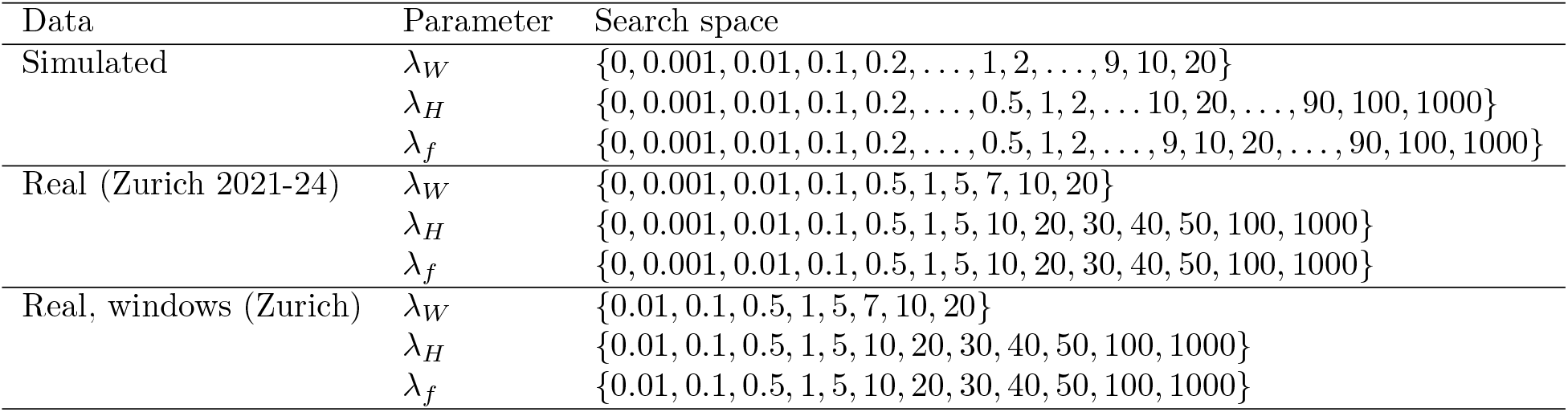
Hyperparameter values tested in 10-fold cross-validation for simulated and real data.

| Data | Parameter | Search space |
|---|---|---|
| Simulated | λ<sub>W</sub> | {0, 0.001, 0.01, 0.1, 0.2, …, 1, 2, …, 9, 10, 20} |
|  | λ<sub>H</sub> | {0, 0.001, 0.01, 0.1, 0.2, …, 0.5, 1, 2, … 10, 20, …, 90, 100, 1000} |
|  | λ<sub>f</sub> | {0, 0.001, 0.01, 0.1, 0.2, …, 0.5, 1, 2, …, 9, 10, 20, …, 90, 100, 1000} |
| Real (Zurich 2021-24) | λ<sub>W</sub> | {0, 0.001, 0.01, 0.1, 0.5, 1, 5, 7, 10, 20} |
|  | λ<sub>H</sub> | {0, 0.001, 0.01, 0.1, 0.5, 1, 5, 10, 20, 30, 40, 50, 100, 1000} |
|  | λ<sub>f</sub> | {0, 0.001, 0.01, 0.1, 0.5, 1, 5, 10, 20, 30, 40, 50, 100, 1000} |
| Real, windows (Zurich) | λ<sub>W</sub> | {0.01, 0.1, 0.5, 1, 5, 7, 10, 20} |
|  | λ<sub>H</sub> | {0.01, 0.1, 0.5, 1, 5, 10, 20, 30, 40, 50, 100, 1000} |
|  | λ<sub>f</sub> | {0.01, 0.1, 0.5, 1, 5, 10, 20, 30, 40, 50, 100, 1000} |

**Table S4:** Fitting settings used for each fold and hyperparameter combination in cross-validation. The solution with the lowest objective function value across random initializations was selected.

| Setting | Simulations | Real data | Real data windows |
|---|---|---|---|
| Random initializations (*n*<sub>run</sub>) | 10 | 5 | 5 |
| Convergence threshold *τ* (relative change of the objective) | 10<sup>−3</sup> | 10<sup>−3</sup> | 10<sup>−3</sup> |
| Maximum number of iterations (*I*<sub>max</sub>) | 5000 | 500 | 500 |
| Binarization target parameter *ρ* | 0 | 0 | 0.1 |

### S2.4 Parameter selection, diagnostics and results from simulated data

**Figure S7:**
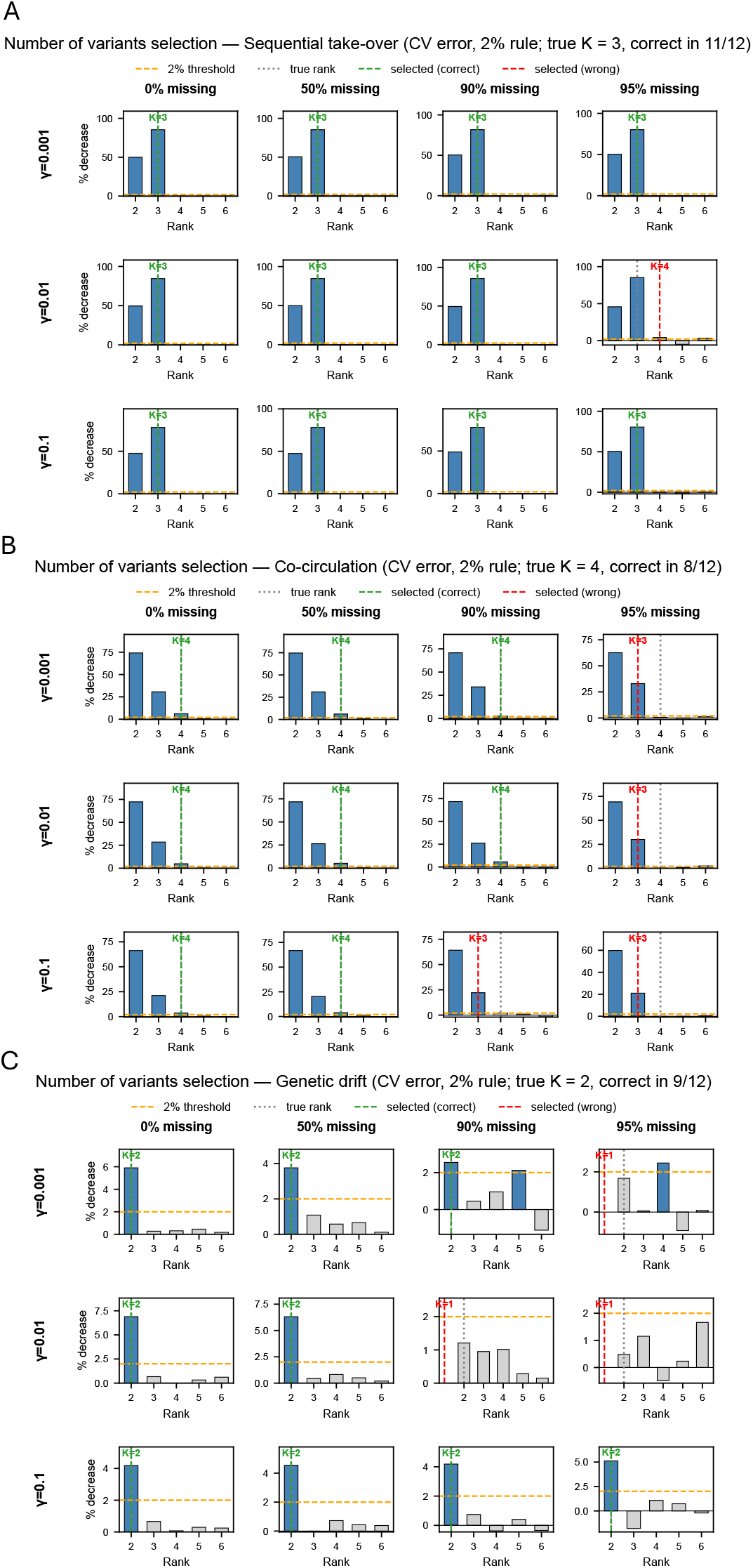
Selection of the number of variants (rank *k*) on simulated data. (A) Three variants with distinct mutation profiles replacing each other sequentially (Section S1.6). Each panel shows relative decrease (%) in held-out cross-validation error between consecutive ranks across the range of candidate variants. The horizontal orange dashed line denotes the threshold of 2% of the relative decrease. Rows: overdispersion *γ* ∈ {0.001, 0.01, 0.1}; columns: fraction of missing values ∈ {0, 50, 90, 95%}. The selected rank (vertical dashed line) is the smallest *k* for which increasing the rank to *k* + 1 reduced the error by less than 2%. (B) same as (A) but for four co-circulating variants of similar mutational profiles (Section S1.6). (C) same as (A) but for two variants evolving under neutral drift (Section S1.6).

**Figure S8:**
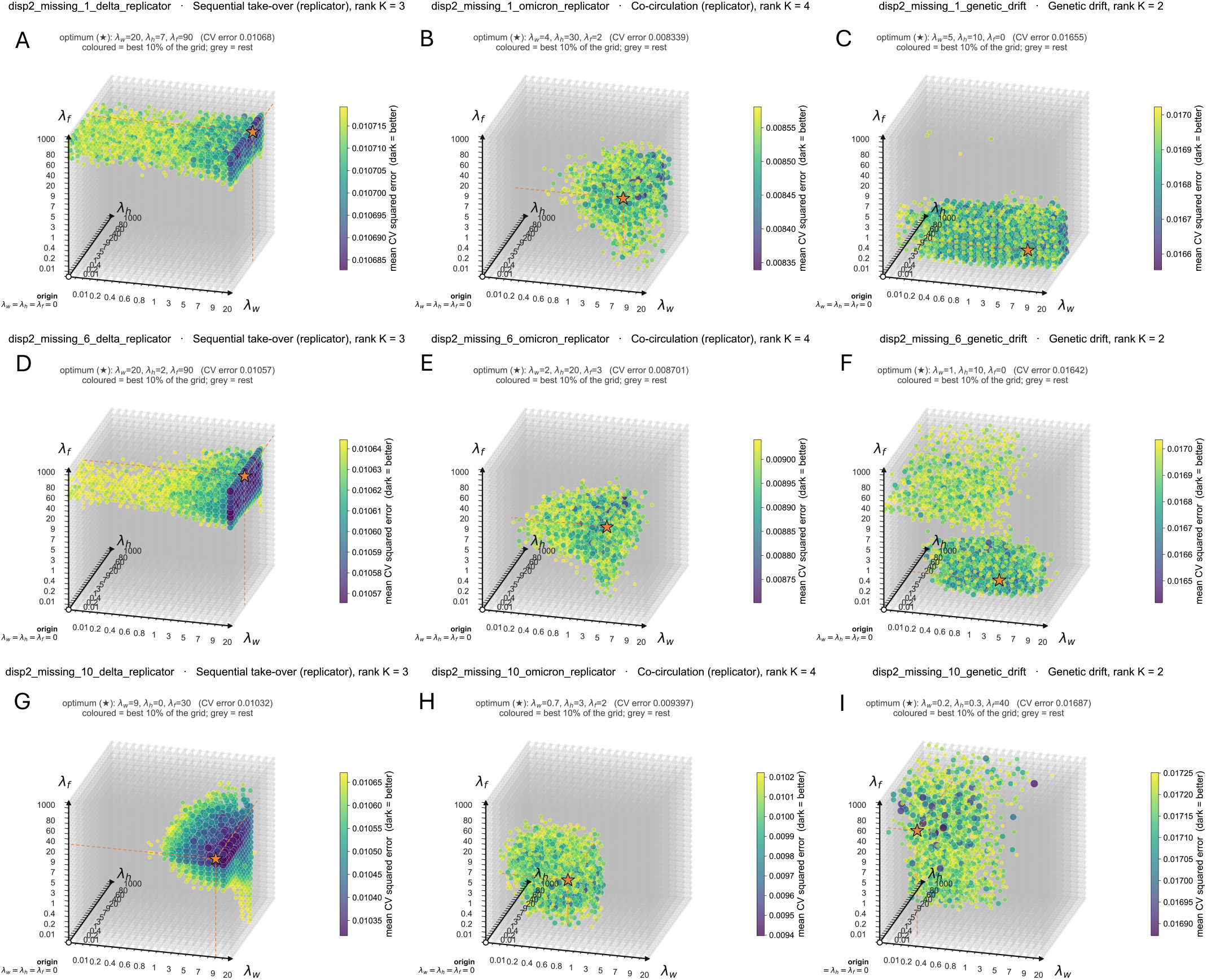
Hyperparameter selection by cross-validation on simulated wastewater data. Columns: three simulated scenarios (Sequential take-over under replicator dynamics, *k* = 3; Co-circulating Omicron-variants under replicator dynamics, *k* = 4; genetic drift, *k* = 2). Rows: 0, 50, 90% missing values; overdispersion *γ* = 0.01 in all panels. (A–I) Mean squared error on held-out entries (averaged over folds) for combinations of *λ*_*W*_, *λ*_*H*_, and *λ*_*f*_ at the true rank *k*. The 10% of parameter combinations with the lowest error are coloured (darker = lower error; the colour scale is specific to each panel); all others are shown in grey. The orange star marks the combination with the lowest error, with dashed lines projecting it onto the axes of corresponding hyperparameter values.

**Figure S9:**
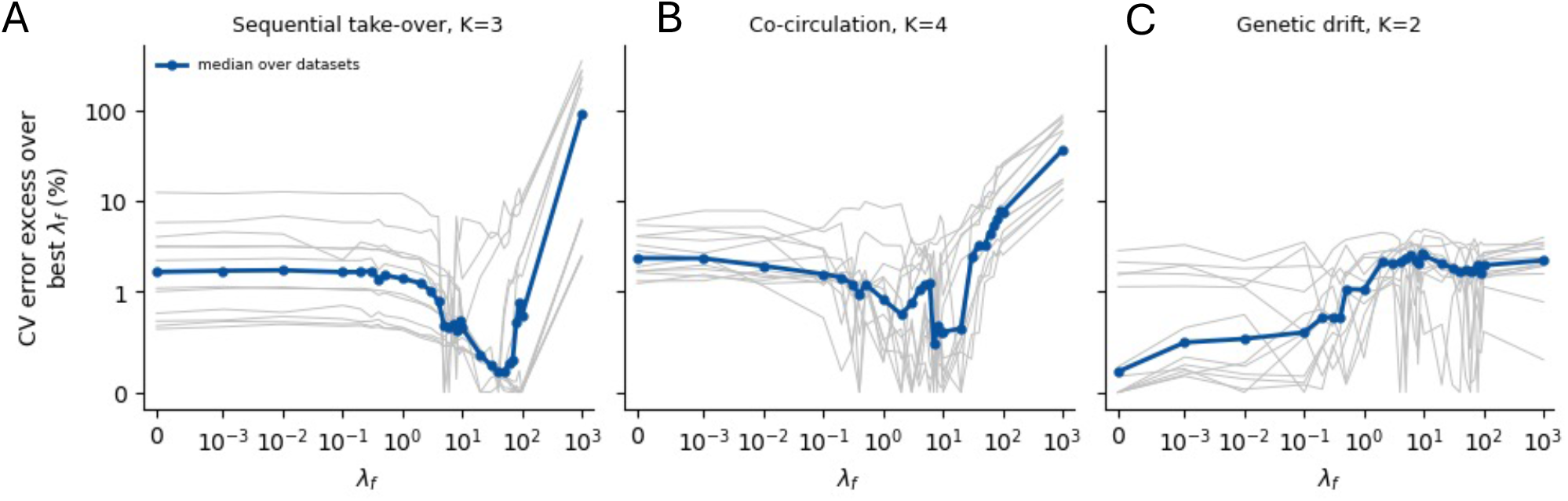
Sensitivity of the cross-validation (CV) error to the fitness regularization *λ*_*f*_ in the simulations across three settings: (A) sequential take-over, (B) co-circulation, and (C) Genetic drift. For each simulated dataset, the CV error at the true number of variants *k* is shown as a function of *λ*_*f*_, where for each *λ*_*f*_ the best combination of *λ*_*W*_ and *λ*_*H*_ was chosen. The y-axis is the increase of this error relative to the lowest error across all *λ*_*f*_ values of the same dataset (0% = best *λ*_*f*_ of that dataset). Thin grey lines show the 12 simulated datasets of each scenario (three overdispersion levels and four fractions of missing data), and the blue line the median across them.

**Figure S10:**
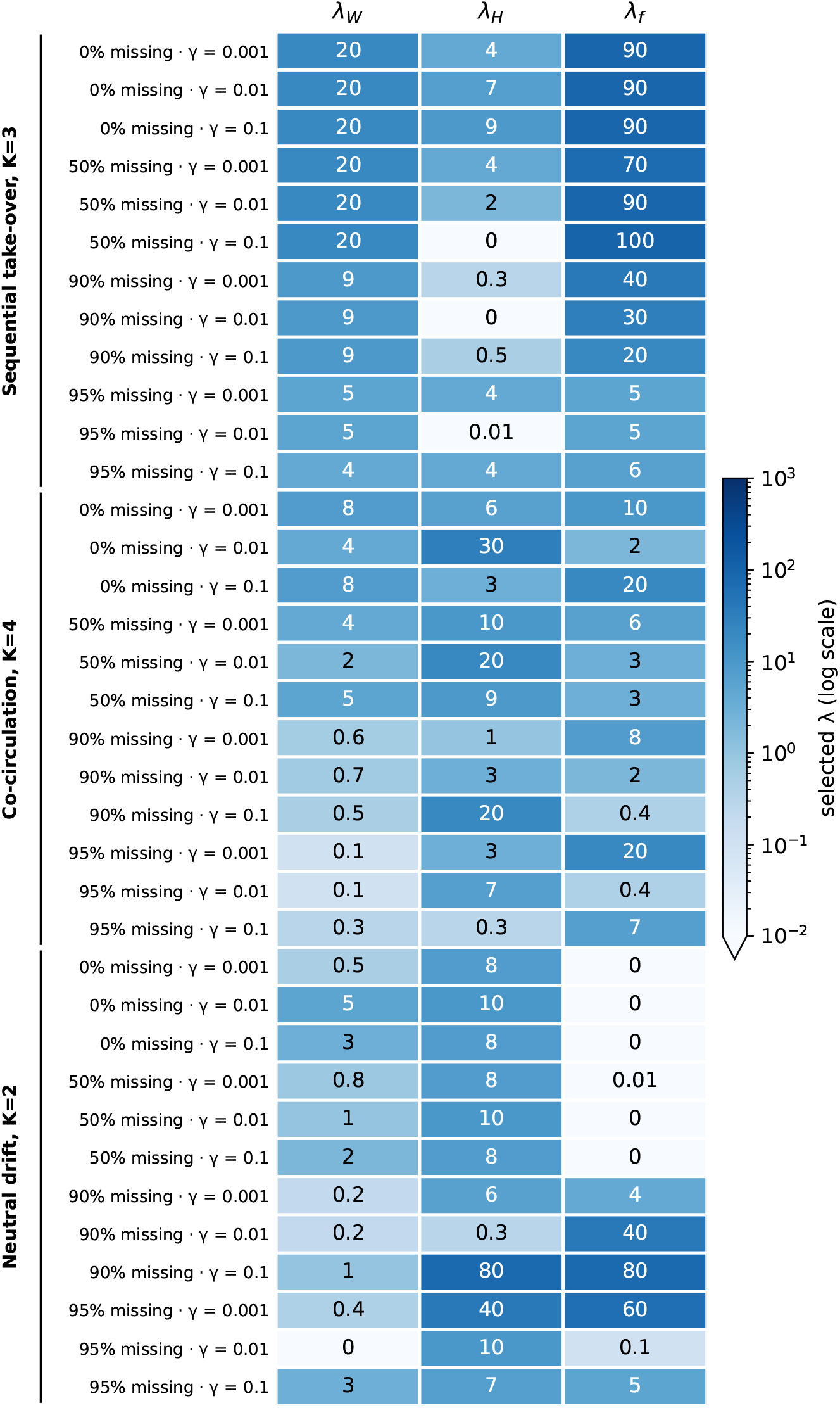
Optimal hyperparameter selected by cross-validation for the three simulated scenarios: sequential take-over (*k* = 3), co-circulation (*k* = 4) and neutral drift, (*k* = 2) at different levels of missing values and overdispersion. For each setting, the hyperparameters (*λ*_*W*_, *λ*_*H*_, *λ*_*f*_) minimizing the cross-validation error at the true rank *k* are shown. In practice, *k* is unknown and must be selected jointly with the hyperparameters over a range of candidate ranks.

**Figure S11:**
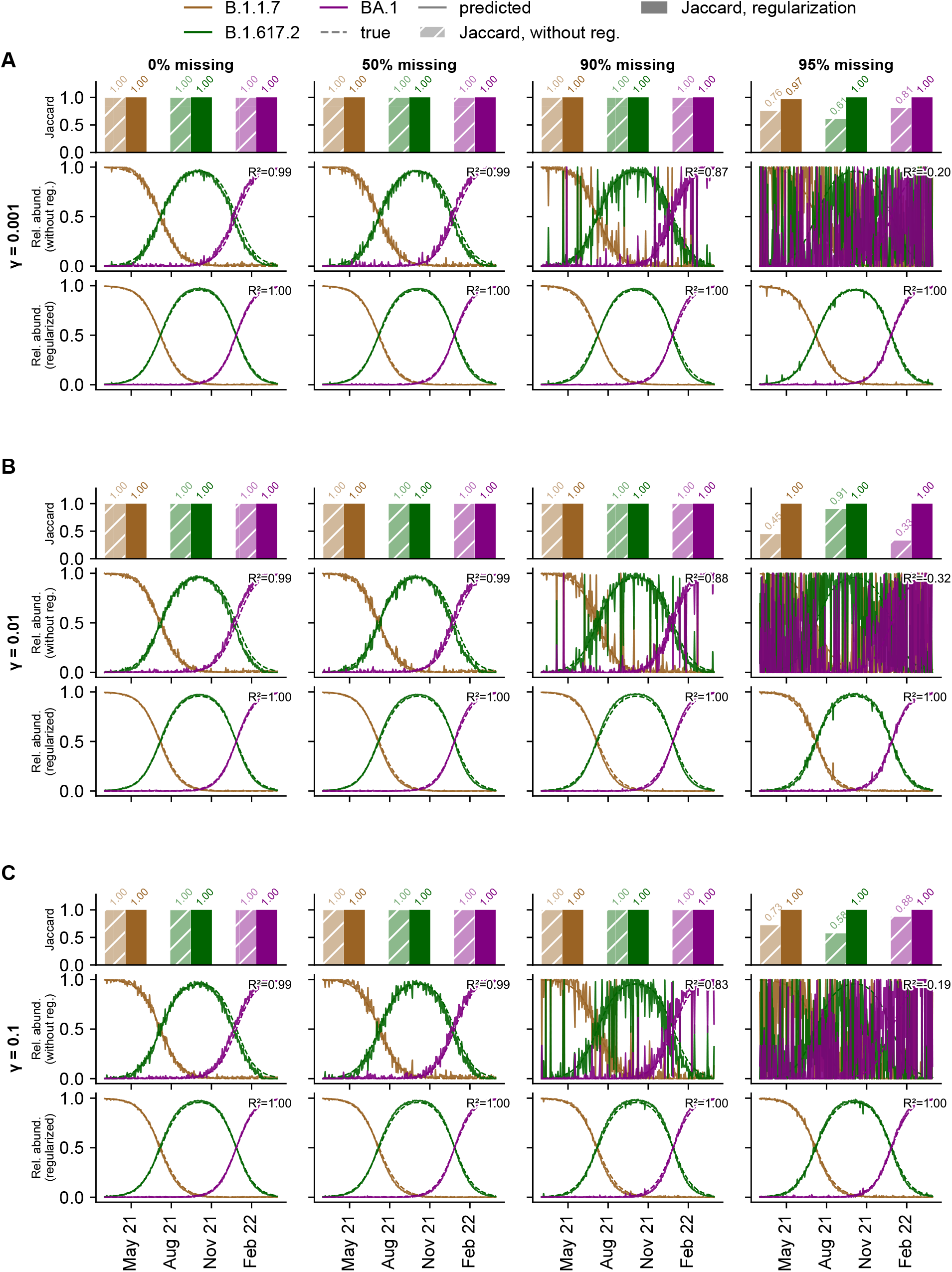
Predicted B.1.1.7, B.1.617.2, and BA.1 variant signatures and relative abundances from simulated mutation frequency data across varying levels of overdispersion and missing values using FitNMF. Predicted trajectories are shown as solid lines and ground-truth values as dashed lines. (A) Low overdispersion (*γ* = 0.001). First row: Jaccard similarity between predicted and true variant signatures for NMF without regularization (*λ*_*H*_ = 0, *λ*_*W*_ = 0, *λ*_*f*_ = 0, hatched bar) and with regularization (hyperparameters selected by cross-validation, solid bar). Second row: predicted relative abundances and *R*^2^ for NMF without regularization. Third row: predicted relative abundances and *R*^2^ for FitNMF. The fraction of missing values increases across columns: 0%, 50%, 90%, and 95%. (B) Same as (A), but with medium overdispersion (*γ* = 0.01). (C) Same as (A), but with high overdispersion (*γ* = 0.1).

**Figure S12:**
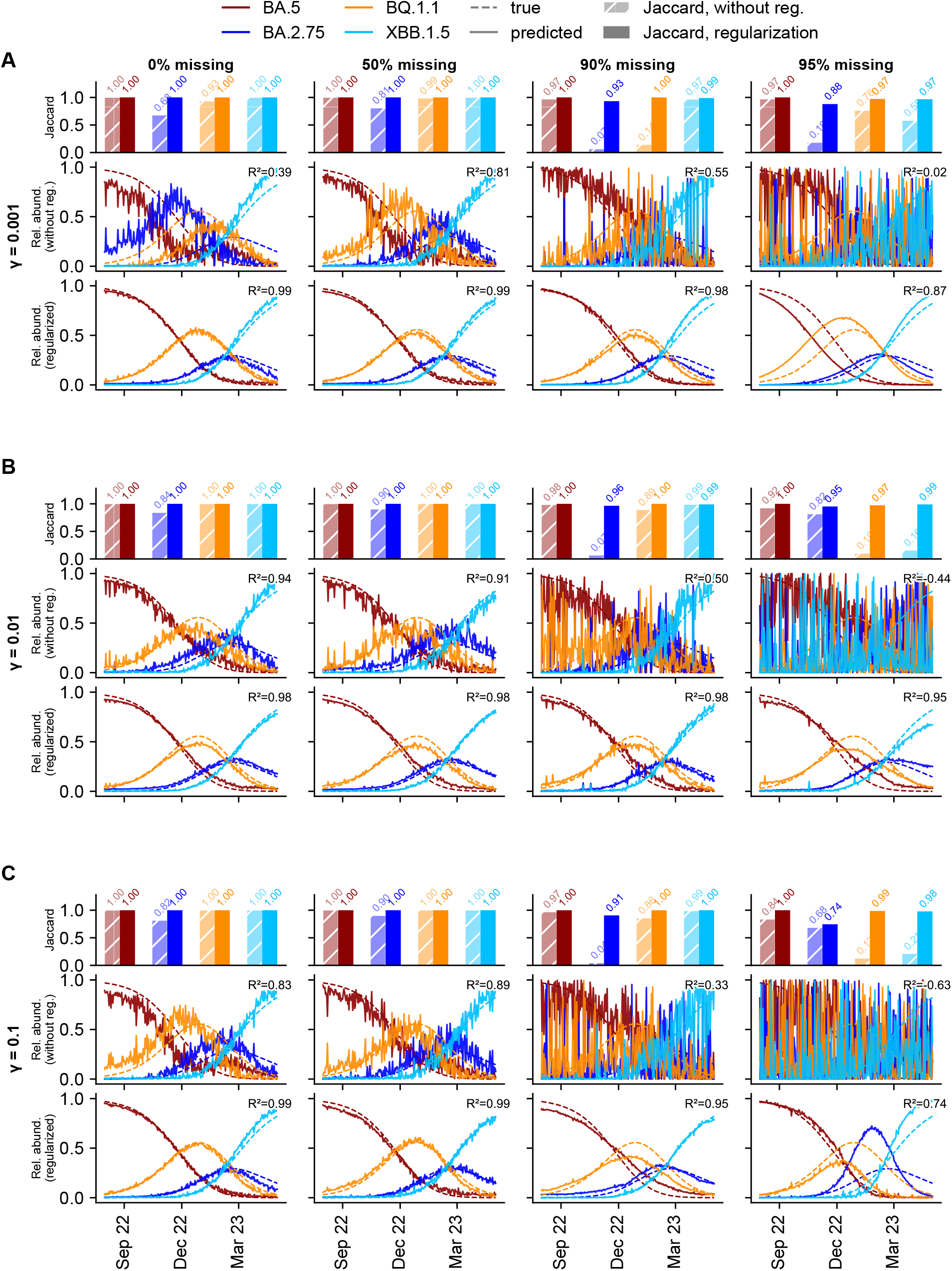
Predicted BA.5, BQ.1.1, BA.2.75, and XBB.1.5 variant signatures and relative abundances from simulated mutation frequency data across varying levels of overdispersion and missing values using FitNMF. Predicted trajectories are shown as solid lines and ground-truth values as dashed lines. (A) Low overdispersion (*γ* = 0.001). First row: Jaccard similarity between predicted and true variant signatures for NMF without regularization (*λ*_*H*_ = 0, *λ*_*W*_ = 0, *λ*_*f*_ = 0, hatched bar) and with regularization (hyperparameters selected by cross-validation, solid bar). Second row: predicted relative abundances and *R*^2^ for NMF without regularization. Third row: predicted relative abundances and *R*^2^ for FitNMF. The fraction of missing values increases across columns: 0%, 50%, 90%, and 95%. (B) Same as (A), but with medium overdispersion (*γ* = 0.01). (C) Same as (A), but with high overdispersion (*γ* = 0.1).

**Figure S13:**
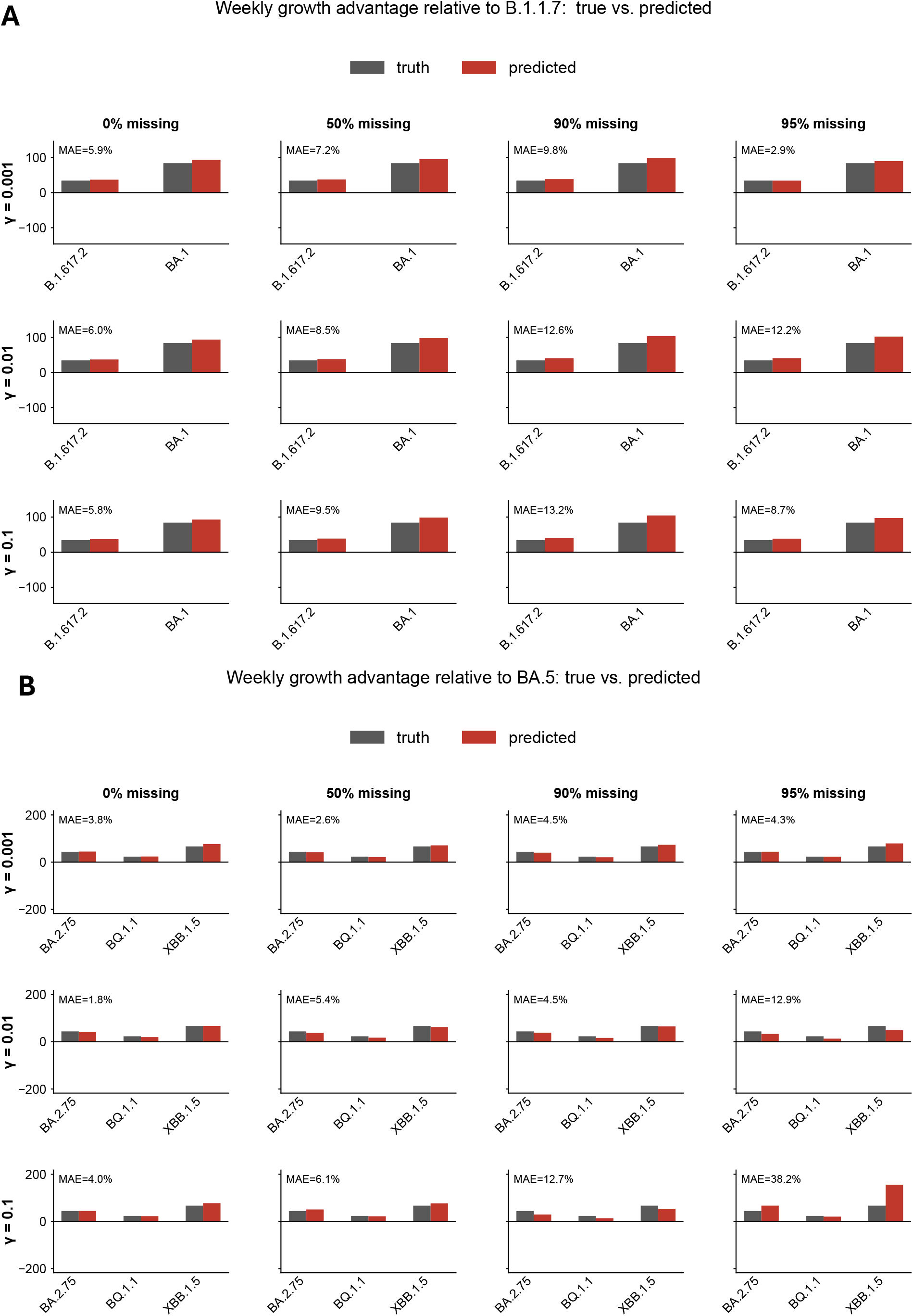
Weekly growth advantage estimates from simulated data using FitNMF. (A) Growth advantages estimated for different SARS-CoV-2 lineages (sequential take-over setting), with B.1.1.7 as reference. (B) Growth advantages estimated for closely related Omicron variants (co-circulation setting), with BA.5 as reference. Growth advantages are expressed as percent change per week. Bars show estimates across simulated datasets with varying overdispersion (rows) and missing-data (columns) levels. Grey bars denote ground-truth growth advantages; red bars denote predicted growth advantages. MAE is the mean absolute error across variants (percentage points).

## S3 Emerging variant detection

**Figure S14:**
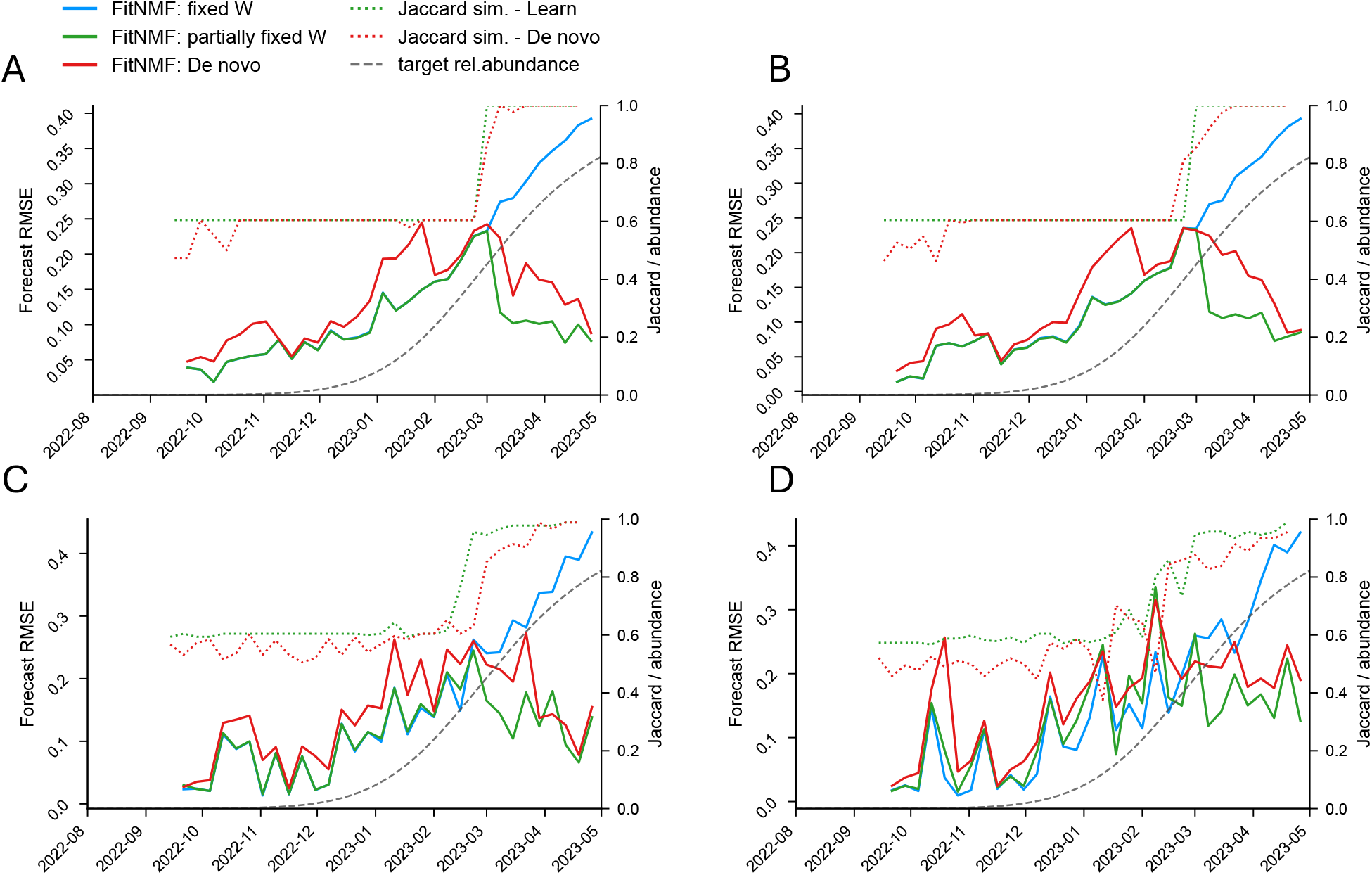
Emerging variant (XBB.1.5) detection using FitNMF on simulated data with (A–D) 0, 50, 90, and 95% missing values, respectively. Left y-axis: forecast RMSE of FitNMF with fixed, known signatures (BA.5, BA.2.75, BQ.1.1; blue), of FitNMF with known signatures fixed plus one unknown component (green), and of FitNMF without prior knowledge of the signatures (*de novo*; red). Each model is trained on all samples up to a given date (training window extended in steps of 7 days) and forecasts the following samples (7 days); RMSE is plotted at the end of this forecast window. Right y-axis: Jaccard similarity between the ground-truth signature of XBB.1.5 and the free component (green, dotted) or the matched *de novo* component (red, dotted), plotted at the end of the training window, and the true relative abundance of XBB.1.5 (grey, dashed).

**Figure S15:**
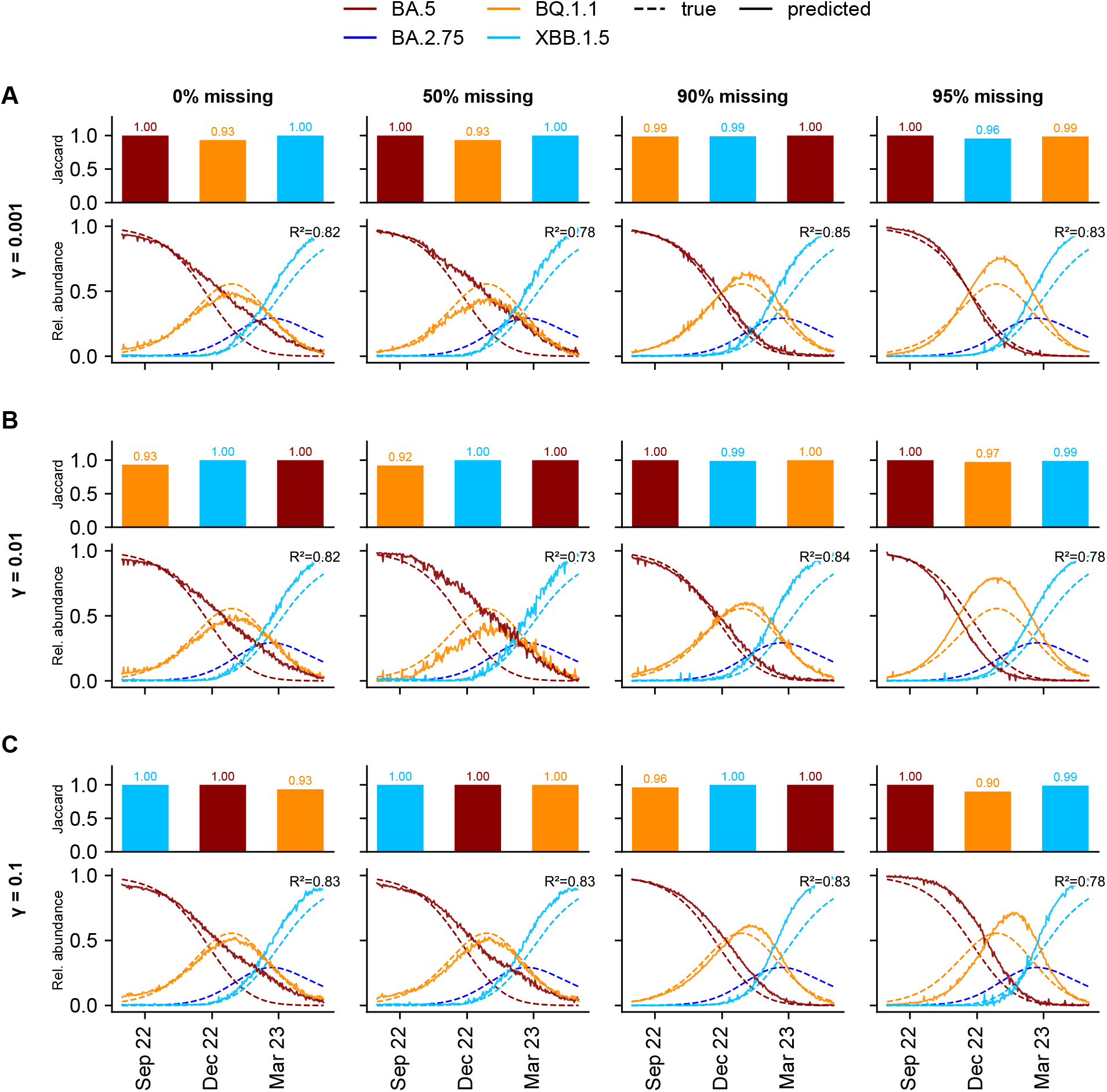
Predicted Omicron variant signatures and relative abundances from simulated mutation frequency data with sub-optimal rank (one too low) across varying levels of overdispersion and missing values using FitNMF method (hyperparameters selected by cross-validation). Predicted trajectories are shown as solid lines and ground-truth values as dashed lines. First row of each subplot: Jaccard similarity between predicted and true variant signatures for FitNMF (A-C). Second row: predicted relative abundances and *R*^2^ for FitNMF (A-C). The true abundance of the variant, whose signature could not be resolved at this rank, was still included in the *R*^2^ evaluation. The fraction of missing values increases across columns: 0%, 50%, 90%, and 95%. The overdispersion levels are indicated on the left.

**Figure S16:**
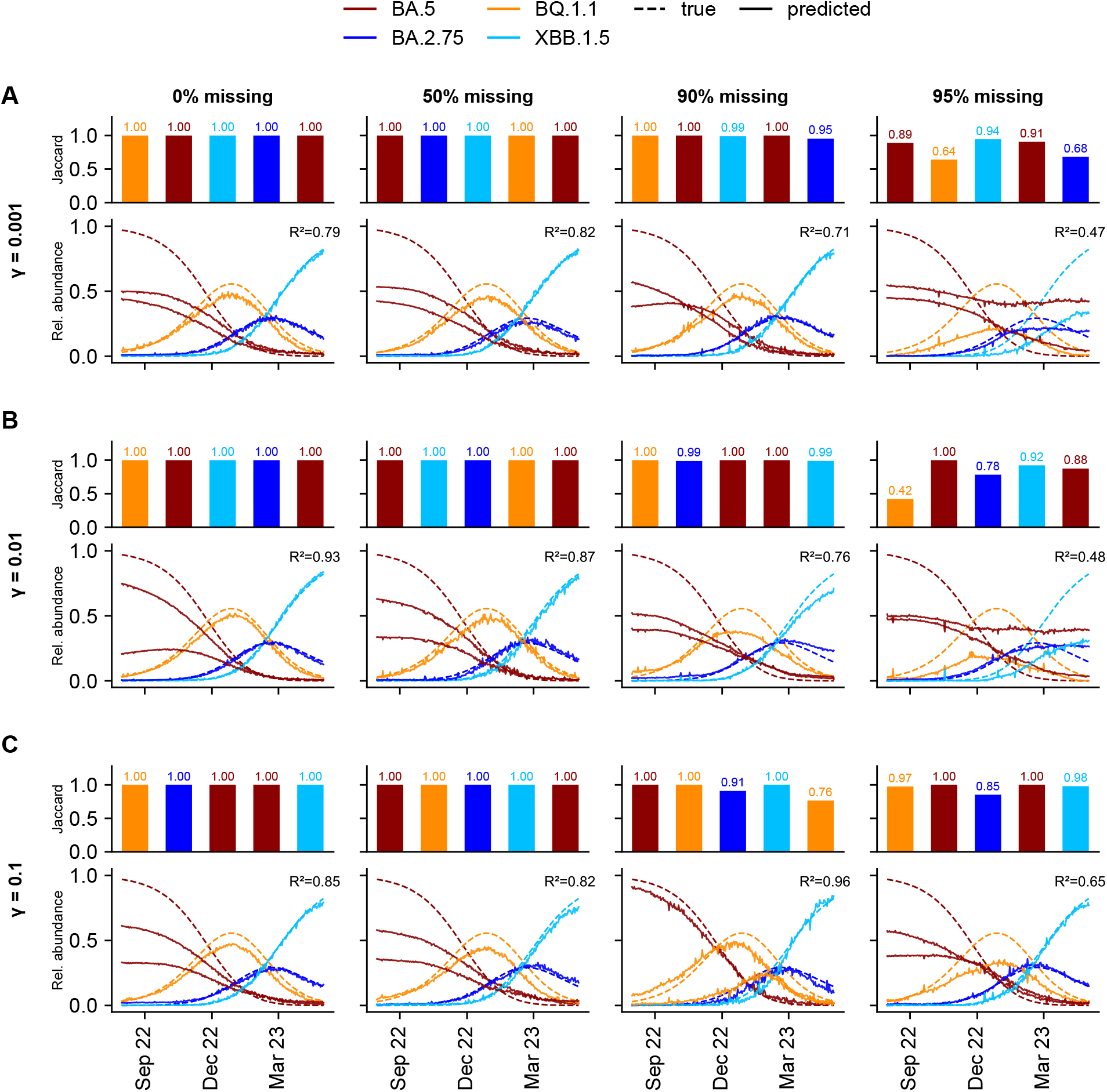
Predicted Omicron variant signatures and relative abundances from simulated mutation frequency data with sub-optimal rank (one too high) across varying levels of overdispersion and missing values using FitNMF (hyperparameters selected by cross-validation). Predicted trajectories are shown as solid lines and ground-truth values as dashed lines. First row of each subplot: Jaccard similarity between predicted and true variant signatures for FitNMF (A-C). Second row: predicted relative abundances and *R*^2^ for FitNMF (A-C). The fraction of missing values increases across columns: 0%, 50%, 90%, and 95%. The overdispersion levels are indicated on the left.

**Figure S17:**
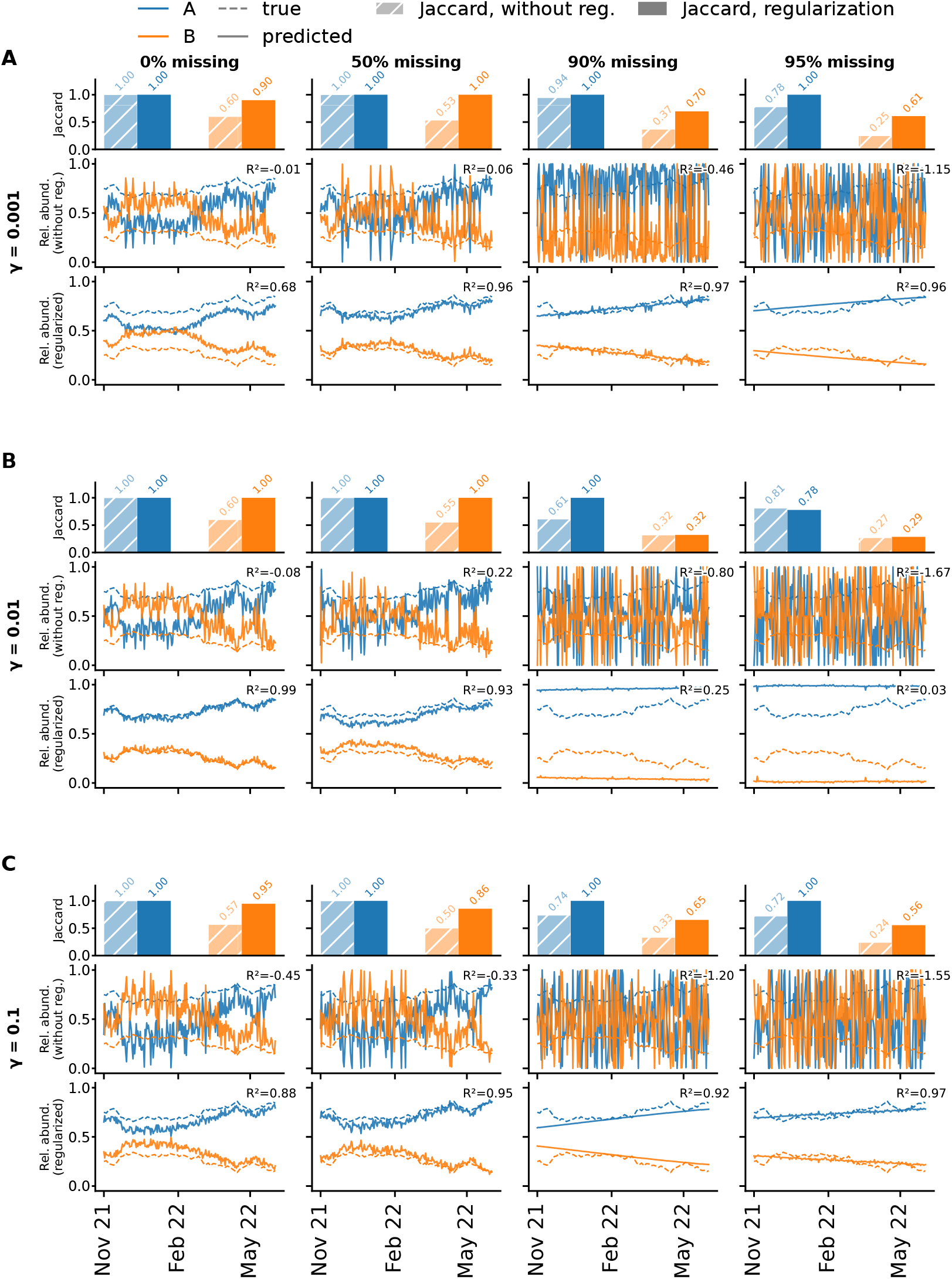
Recovery of variant signatures and relative abundances by FitNMF on simulated mutation-frequency data with two variants whose abundances follow genetic drift (no selective advantage; Section S1.6), across levels of overdispersion *γ* and fractions of missing values. Predicted trajectories are shown as solid lines and ground truth as dashed lines. (A) Low overdispersion (*γ* = 0.001). First row: Jaccard similarity between predicted and true variant signatures for unregularized NMF (*λ*_*W*_ = *λ*_*H*_ = *λ*_*f*_ = 0, hatched bars) and for NMF with regularization (hyperparameters selected by cross-validation, solid bars). Second row: predicted relative abundances and *R*^2^ for unregularized NMF. Third row: predicted relative abundances and *R*^2^ for FitNMF. The fraction of missing values increases across columns: 0%, 50%, 90%, and 95%. (B) As (A), with medium overdispersion (*γ* = 0.01). (C) As (A), with high overdispersion (*γ* = 0.1).

### S3.1 Parameter selection, diagnostics and results from real data

**Figure S18:**
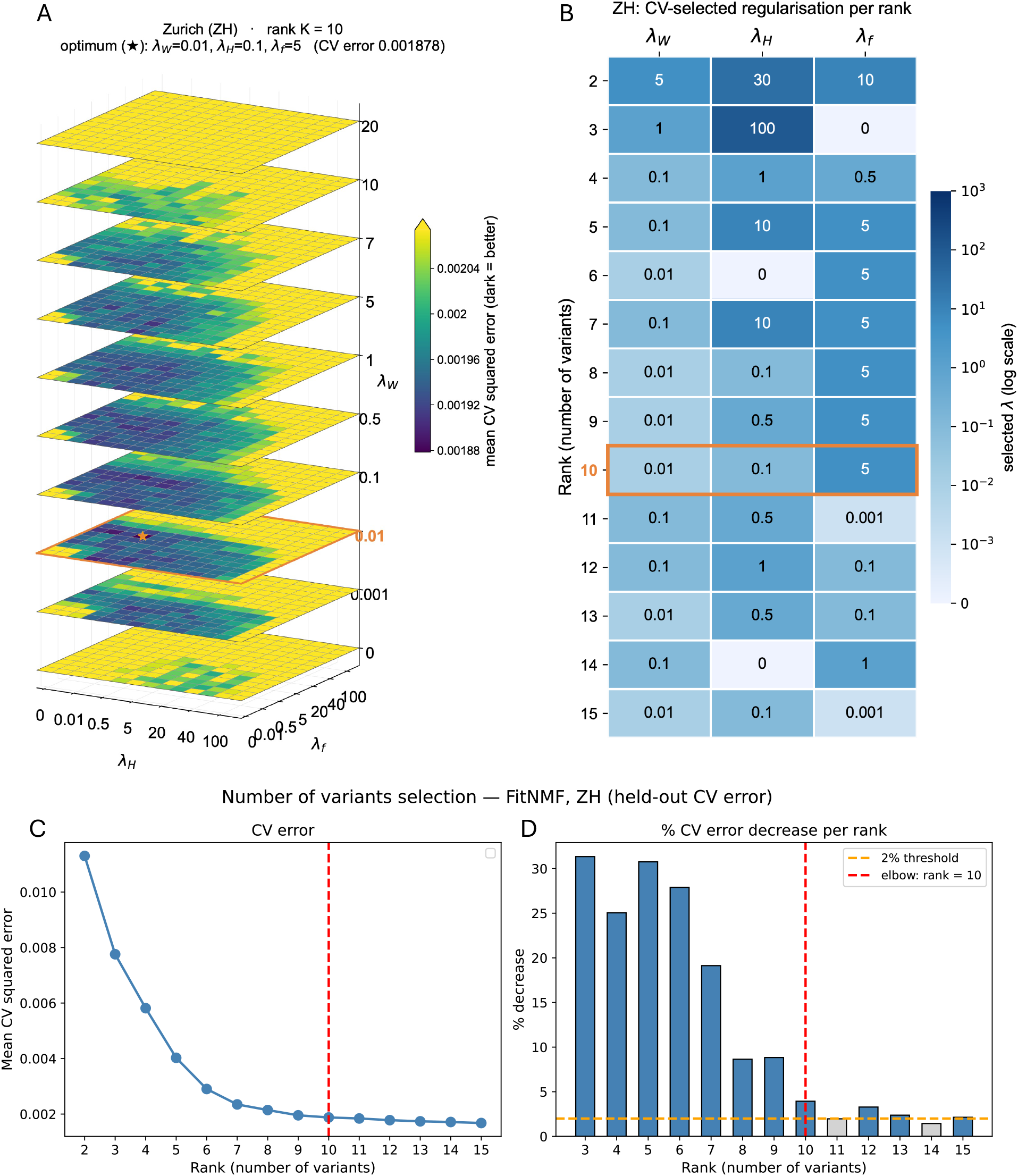
Hyperparameter and rank selection by cross-validation on wastewater data from the Zurich WWTP, 2021–2024. In each of 10 folds, 10% of the observed entries of the mutation-frequency matrix were held out, and the model was fit on the remaining entries. (A) Mean squared error on held-out entries (averaged over folds) for combinations of *λ*_*W*_, *λ*_*H*_ and *λ*_*f*_ at rank *k* = 10. Darker shades indicate lower error; the colour scale spans the minimum to the median of all errors. (B) Optimal hyperparameters for each rank *k* = 2, …, 15. (C) Mean held-out squared error as a function of rank *k*. (D) Relative decrease in held-out error between consecutive ranks. The selected rank *k* (red vertical dashed line) is the smallest *k* for which increasing the rank to *k* + 1 reduced the error by less than 2% (orange horizontal dashed line).

**Figure S19:**
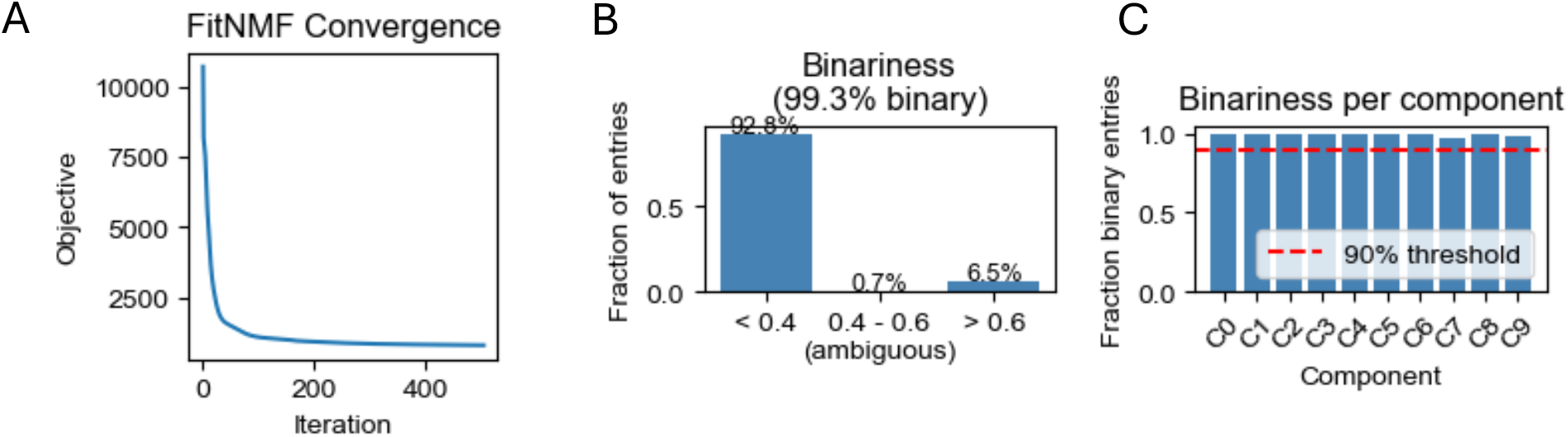
Diagnostic plots of the FitNMF fit on wastewater sequencing data from Zurich (ZH) WWTP, with rank *k* = 10. (A) Objective function value over iterations for the best of 10 random restarts. (B) Percentage of entries of the estimated signature matrix *W* falling into the ranges *<* 0.4, [0.4, 0.6), and ≥ 0.6 across components. (C) Fraction of non-ambiguous entries (≤ 0.4 or ≥ 0.6) per component, *k* = 1, …, 10.

**Figure S20:**
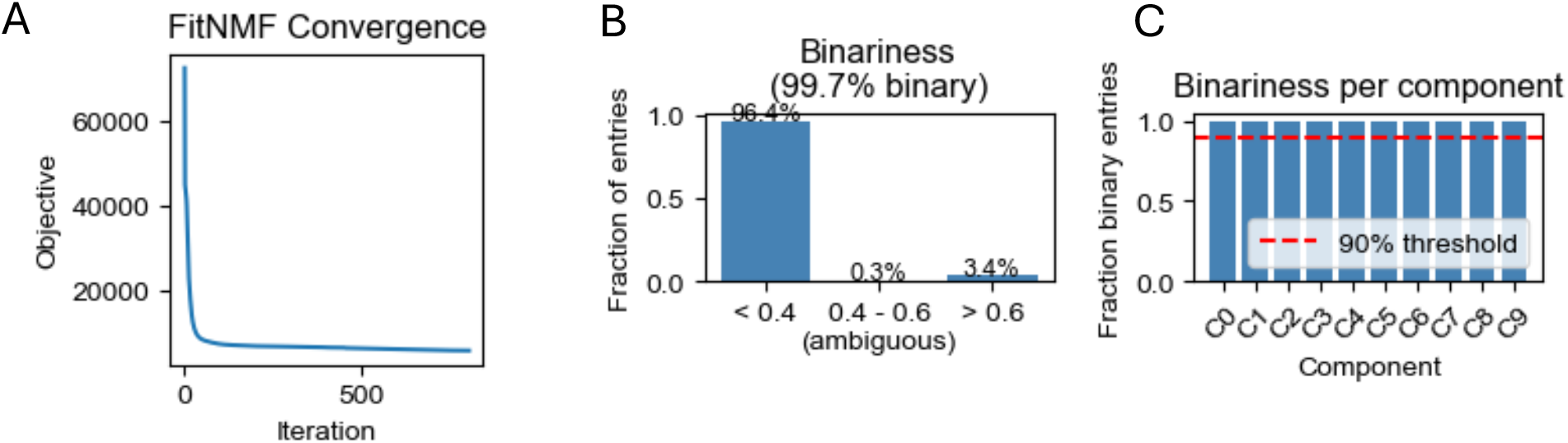
Diagnostic plots of the FitNMF fit on wastewater sequencing data from six Swiss WWTPs (Zurich (ZH), Chur (GR), Laupen (BE), Geneva (GE), Altenrhein (SG), Lugano (TI)) jointly, with rank *k* = 10. (A) Objective function value over iterations for the best of 10 random restarts. (B) Percentage of entries of the estimated signature matrix *W* falling into the ranges *<* 0.4, [0.4, 0.6), and ≥ 0.6 across components. (C) Fraction of non-ambiguous entries (≤ 0.4 or ≥ 0.6) per component, *k* = 1, …, 10.

## References

[1] W. Ahmed, N. Angel, J. Edson, K. Bibby, A. Bivins, J. W. O’Brien, P. M. Choi, M. Kitajima, S. L. Simpson, J. Li, B. Tscharke, R. Verhagen, W. J. Smith, J. Zaugg, L. Dierens, P. Hugenholtz, K. V. Thomas, and J. F. Mueller, “First confirmed detection of sars-cov-2 in untreated wastewater in australia: A proof of concept for the wastewater surveillance of covid-19 in the community,” Science of The Total Environment, vol. 728, p. 138764, 2020. [Online]. Available: https://www.sciencedirect.com/science/article/pii/S0048969720322816

[2] J. S. Huisman, J. Scire, L. Caduff, X. Fernandez-Cassi, P. Ganesanandamoorthy, A. Kull, A. Scheidegger, E. Stachler, A. B. Boehm, B. Hughes, A. Knudson, A. Topol, K. R. Wigginton, M. K. Wolfe, T. Kohn, C. Ort, T. Stadler, and T. R. Julian, “Wastewater-based estimation of the effective reproductive number of sars-cov-2,” Environmental Health Perspectives, vol. 130, no. 5, p. 057011, 05 2022. [Online]. Available: 10.1289/EHP10050

[3] S. Karthikeyan, J. I. Levy, P. De Hoff, G. Humphrey, A. Birmingham, K. Jepsen, S. Farmer, H. M. Tubb, T. Valles, C. E. Tribelhorn, R. Tsai, S. Aigner, S. Sathe, N. Moshiri, B. Henson, A. M. Mark, A. Hakim, N. A. Baer, T. Barber, P. Belda-Ferre, M. Chacón, W. Cheung, E. S. Cresini, E. R. Eisner, A. L. Lastrella, E. S. Lawrence, C. A. Marotz, T. T. Ngo, T. Ostrander, A. Plascencia, R. A. Salido, P. Seaver, E. W. Smoot, D. McDonald, R. M. Neuhard, A. L. Scioscia, A. M. Satterlund, E. H. Simmons, D. B. Abelman, D. Brenner, J. C. Bruner, A. Buckley, M. Ellison, J. Gattas, S. L. Gonias, M. Hale, F. Hawkins, L. Ikeda, H. Jhaveri, T. Johnson, V. Kellen, B. Kremer, G. Matthews, R. W. McLawhon, P. Ouillet, D. Park, A. Pradenas, S. Reed, L. Riggs, A. Sanders, B. Sollenberger, A. Song, B. White, T. Winbush, C. M. Aceves, C. Anderson, K. Gangavarapu, E. Hufbauer, E. Kurzban, J. Lee, N. L. Matteson, E. Parker, S. A. Perkins, K. S. Ramesh, R. Robles-Sikisaka, M. A. Schwab, E. Spencer, S. Wohl, L. Nicholson, I. H. McHardy, D. P. Dimmock, C. A. Hobbs, O. Bakhtar, A. Harding, A. Mendoza, A. Bolze, D. Becker, E. T. Cirulli, M. Isaksson, K. M. Schiabor Barrett, N. L. Washington, J. D. Malone, A. M. Schafer, N. Gurfield, S. Stous, R. Fielding-Miller, R. S. Garfein, T. Gaines, C. Anderson, N. K. Martin, R. Schooley, B. Austin, D. R. MacCannell, S. F. Kingsmore, W. Lee, S. Shah, E. McDonald, A. T. Yu, M. Zeller, K. M. Fisch, C. Longhurst, P. Maysent, D. Pride, P. K. Khosla, L. C. Laurent, G. W. Yeo, K. G. Andersen, and R. Knight, “Wastewater sequencing reveals early cryptic sars-cov-2 variant transmission,” Nature, vol. 609, no. 7925, pp. 101–108, 2022.

[4] D. Dreifuss, I. Topolsky, P. Icer Baykal, and N. Beerenwinkel, “Tracking sars-cov-2 genomic variants in wastewater sequencing data with lollipop,” PLOS Computational Biology, vol. 22, pp. 1–19, 02 2026. [Online]. Available: 10.1371/journal.pcbi.1014003

[5] S. Edmunds, M. Ajelli, M. Litvinova, and D. Landsittel, “Comparing wastewater-based and case-based r(t) estimates of sars-cov-2 transmission in georgia using generalized linear mixed models,” Epidemiology and Infection, vol. 154, pp. 1–19, 2026. [Online]. Available: 10.1017/S0950268826101356

[6] E. Volz, “Fitness, growth and transmissibility of sars-cov-2 genetic variants,” Nature Reviews Genetics, vol. 24, no. 10, pp. 724–734, 2023.

[7] W. Boulton, F. R. Fidan, H. Denise, N. De Maio, and N. Goldman, “Swampy: simulating sars-cov-2 wastewater amplicon metagenomes,” Bioinformatics, vol. 40, no. 9, p. btae532. 09 2024. [Online]. Available: 10.1093/bioinformatics/btae532

[8] C. Brown, J. Vostok, H. Johnson, M. Burns, R. Gharpure, S. Sami, R. Sabo, N. Hall, A. Foreman, P. Schubert, G. Gallagher, T. Fink, L. Madoff, S. Gabriel, B. MacInnis, D. Park, K. Siddle, V. Harik, D. Arvidson, T. Brock-Fisher, M. Dunn, A. Kearns, and A. Laney, “Outbreak of sars-cov-2 infections, including covid-19 vaccine breakthrough infections, associated with large public gatherings – barnstable county, massachusetts, july 2021,” MMWR Morbidity and Mortality Weekly Report, vol. 70, no. 31, pp. 1059–1062, aug 2021.

[9] J. de Korne-Elenbaas, A. Rimaite, I. Topolsky, D. Dreifuss, C. Bürki, L. Fuhrmann, L. du Plessis, W. J. Fitzsimmons, E. E. Bendall, T. Stadler, N. Beerenwinkel, and T. R. Julian, “Wastewater-based sequencing of respiratory syncytial virus to investigate lineage dynamics and antigenic site mutations: a retrospective genomic epidemiology study,” The Lancet Microbe, 2026. [Online]. Available: 10.1016/j.lanmic.2026.101355

[10] A. Rimaite, J. de Korne-Elenbaas, A. Lison, T. Stadler, T. R. Julian, and N. Beerenwinkel, “Tracking respiratory syncytial virus dynamics in wastewater during the 2024-2025 season in switzerland,” medRxiv, pp. 2026–05, 2026.

[11] A. John, S. Kang, L. Fuhrmann, I. Topolsky, C. Kent, J. Quick, T. Stadler, T. R. Julian, and N. Beerenwinkel, “Characterizing influenza a virus lineages and clinically relevant mutations through high-coverage wastewater sequencing,” Water Research, vol. 287, no. Pt B, p. 124453, Aug. 2025, epub 2025 Aug 21. [Online]. Available: 10.1016/j.watres.2025.124453

[12] G. Medema, L. Heijnen, G. Elsinga, R. Italiaander, and A. Brouwer, “Presence of sars-coronavirus-2 rna in sewage and correlation with reported covid-19 prevalence in the early stage of the epidemic in the netherlands,” Environmental Science & Technology Letters, vol. 7, no. 7, pp. 511–516, 2020. [Online]. Available: 10.1021/acs.estlett.0c00357

[13] M. Pitton, R. E. McLeod, L. Caduff, A. Dauletova, J. de Korne-Elenbaas, C. Gan, C. Hablützel, A. Holschneider, S. Kang, G. Loustalot et al., “A six-plex digital pcr assay for monitoring respiratory viruses in wastewater,” Nature Water, vol. 3, no. 10, pp. 1174–1186, 2025.

[14] K. Jahn, D. Dreifuss, I. Topolsky, A. Kull, P. Ganesanandamoorthy, X. Fernandez-Cassi, C. Bänziger, A. J. Devaux, E. Stachler, L. Caduff, F. Cariti, A. T. Corzón, L. Fuhrmann, C. Chen, K. P. Jablonski, S. Nadeau, M. Feldkamp, C. Beisel, C. Aquino, T. Stadler, C. Ort, T. Kohn, T. R. Julian, and N. Beerenwinkel, “Early detection and surveillance of sars-cov-2 genomic variants in wastewater using cojac,” Nature Microbiology, vol. 7, no. 8, pp. 1151–1160, 2022.

[15] D. Dreifuss, P. Czyż, and N. Beerenwinkel, “Learning and forecasting selection dynamics of sars-cov-2 variants from wastewater sequencing data using covvfit,” Water Research, vol. 301, p. 126018, 2026. [Online]. Available: https://www.sciencedirect.com/science/article/pii/S0043135426006998

[16] P. V. Markov, M. Ghafari, M. Beer, K. Lythgoe, P. Simmonds, N. I. Stilianakis, and A. Katzourakis, “The evolution of sars-cov-2,” Nature Reviews Microbiology, vol. 21, no. 6, pp. 361–379, 2023.

[17] Y. Liu, N. Sapoval, P. Gallego-García, L. Tomás, D. Posada, T. J. Treangen, and L. B. Stadler, “Crykey: rapid identification of sars-cov-2 cryptic mutations in wastewater,” Nature Communications, vol. 15, no. 1, p. 4545, 2024.

[18] R. Suarez, D. A. Gregory, D. A. Baker, C. A. Rushford, T. L. Hunter, N. R. Minor, C. M. Russ, E. E. Copen, D. H. O’Connor, and M. C. Johnson, “Detecting sars-cov-2 cryptic lineages using publicly available whole genome wastewater sequencing data,” PLOS Pathogens, vol. 21, no. 6, pp. 1–15, 2025. [Online]. Available: 10.1371/journal.ppat.1012850

[19] F. Amman, R. Markt, L. Endler, S. Hupfauf, B. Agerer, A. Schedl, L. Richter, M. Zechmeister, M. Bicher, G. Heiler, P. Triska, M. Thornton, T. Penz, M. Senekowitsch, J. Laine, Z. Keszei, P. Klimek, F. Nägele, M. Mayr, B. Daleiden, M. Steinlechner, H. Niederstätter, P. Heidinger, W. Rauch, C. Scheffknecht, G. Vogl, G. Weichlinger, A. O. Wagner, K. Slipko, A. Masseron, E. Radu, F. Allerberger, N. Popper, C. Bock, D. Schmid, H. Oberacher, N. Kreuzinger, H. Insam, and A. Bergthaler, “Viral variant-resolved wastewater surveillance of sars-cov-2 at national scale,” Nature Biotechnology, vol. 40, no. 12, pp. 1814–1822, 2022.

[20] A. Lefebvre, V. Maréchal, A. Gloaguen, T. O. Consortium, A. Lambert, and Y. Maday, “Unsupervised detection and fitness estimation of emerging sars-cov-2 variants: Application to wastewater samples (anrs0160),” PLOS Computational Biology, vol. 21, pp. 1–22, 12 2025. [Online]. Available: 10.1371/journal.pcbi.1013749

[21] X. Zhuang, V. Vo, M. A. Moshi, K. Dhede, N. Ghani, S. Akbar, C.-L. Chang, A. K. Young, E. Buttery, W. Bendik, H. Zhang, S. Afzal, D. Moser, D. Cordes, C. Lockett, D. Gerrity, H.-Y. Kan, and E. C. Oh, “Early detection of emerging sars-cov-2 variants from wastewater through genome sequencing and machine learning,” Nature Communications, vol. 16, no. 1, p. 6272, 2025.

[22] I. Ellmen, A. K. Overton, J. J. Knapp, D. Nash, H. Ho, Y. Hungwe, S. Prasla, J. I. Nissimov, and T. C. Charles, “Reconstructing sars-cov-2 lineages from mixed wastewater sequencing data,” Scientific Reports, vol. 14, no. 1, p. 20273, 2024.

[23] D. Becker, “Unmuted: Defining sars-cov-2 lineages according to temporally consistent mutation clusters in wastewater samples,” Epidemics, vol. 54, p. 100876, 2026. [Online]. Available: https://www.sciencedirect.com/science/article/pii/S1755436525000647

[24] M. A. Nowak, Evolutionary dynamics: exploring the equations of life. Cambridge, MA: Harvard University Press, 2006.

[25] D. D. Lee and H. S. Seung, “Algorithms for non-negative matrix factorization,” in Proceedings of the 14th International Conference on Neural Information Processing Systems, ser. NIPS’00. Cambridge, MA, USA: MIT Press, 2000, p. 535–541.

[26] L. Fuhrmann, K. P. Jablonski, I. Topolsky, A. A. Batavia, N. Borgsmüller, P. I. Baykal, M. Carrara, C. Chen, A. Dondi, M. Dragan, D. Dreifuss, A. John, B. Langer, M. Okoniewski, L. du Plessis, U. Schmitt, F. Singer, T. Stadler, and N. Beerenwinkel, “V-pipe 3.0: a sustainable pipeline for within-sample viral genetic diversity estimation,” GigaScience, vol. 13, p. giae065, 2024. [Online]. Available: 10.1093/gigascience/giae065

[27] C. Chen, S. Nadeau, M. Yared, P. Voinov, N. Xie, C. Roemer, and T. Stadler, “Cov-spectrum: analysis of globally shared sars-cov-2 data to identify and characterize new variants,” Bioinformatics, vol. 38, no. 6, pp. 1735–1737, Mar. 2022. [Online]. Available: 10.1093/bioinformatics/btab856

[28] E. Abousamra, M. Figgins, and T. Bedford, “Fitness models provide accurate short-term forecasts of sars-cov-2 variant frequency,” PLOS Computational Biology, vol. 20, no. 9, pp. 1–20, 09 2024. [Online]. Available: 10.1371/journal.pcbi.1012443

[29] R. H. Byrd, P. Lu, J. Nocedal, and C. Zhu, “A limited memory algorithm for bound constrained optimization,” SIAM Journal on Scientific Computing, vol. 16, no. 5, pp. 1190–1208, 1995. [Online]. Available: 10.1137/0916069

[30] H. Li and R. Durbin, “Fast and accurate short read alignment with burrows–wheeler transform,” Bioinformatics, vol. 25, no. 14, pp. 1754–1760, 07 2009. [Online]. Available: 10.1093/bioinformatics/btp324

